# Real-world adherence to guidelines in acute severe ulcerative colitis: automated assessment using large language models

**DOI:** 10.64898/2026.11.18.26368431

**Authors:** Ann-Kathrin Schalkamp, Mao-Yuan Chen, Aryan Ayati, Shadera Azzam, Banaz Shwan, Sara Lewin, Uma Mahadevan, Vivek A. Rudrapatna

**Author notes:** Co-senior authors (Uma Mahadevan, Vivek A. Rudrapatna). Corresponding author: Vivek Ashok Rudrapatna, 490 Illinois Street, San Francisco CA 94158, Fax: N/A. Author contact details: Ann-Kathrin Schalkamp, Mao-Yuan Chen, Aryan Ayati, Shadera Azzam, Banaz Shwan, Sara Lewin, Uma Mahadevan. **Conflict of interest statement** VAR’s research is partially funded by grants to UCSF from the following for-profit entities: Merck, Genentech, Johnson & Johnson, Sanofi, and Blueprint Medicines. He is an advisor to Data Unite and ZebraMD. The authors declare no actual competing interests.

## Abstract

**Background and Aims:** Acute severe ulcerative colitis (ASUC) is a medical emergency which requires close guideline adherence for optimal outcomes. However, measuring real-world adherence to guidelines and identifying guideline deviations are labor-intensive. We assessed how well a large language model (LLM)-based framework can measure real-world adherence to ASUC guidelines and identify reasons for deviations.

**Methods:** The retrospective study focused on clinician adherence to the 2025 American College of Gastroenterology guidelines on ASUC. A multi-agent GPT-5 framework first reviewed inpatient health records to confirm ASUC status. Then for each guideline, it assessed whether it was applicable, recommended by consultants, ordered, or executed. This system was benchmarked against manual chart review. The framework was then applied to all the first seven days of all ASUC hospitalizations in 2019–2024 at UCSF. An additional LLM assessed clinical justification of guideline deviations.

**Results:** The system reliably recognized ASUC hospitalizations (sensitivity: 0.93) and annotated the guideline cascade (overall accuracy: 0.95, N=60). Across 121 hospitalizations (104 patients), adherence was >80%. Deviations concentrated in deep vein thrombosis prophylaxis (DVTp) and timely rescue therapy. Particularly, DVTp broke down at execution despite appropriate recommendation and ordering, whereas "avoidance" recommendations were followed in practice but seldom explicitly documented. Overall, 62% of non-adherence was unjustified, mostly process delays in DVTp.

**Conclusions:** Real-world ASUC care at a tertiary center was largely guideline-concordant, with actionable gaps in DVT prophylaxis execution and rescue therapy timing. A guideline-constrained LLM system enables scalable monitoring of clinical quality and creates the potential in simultaneously improving clinical care and guidelines.

## Introduction

Acute severe ulcerative colitis (ASUC) is a medical emergency associated with substantial morbidity and mortality if evidence-based care is delayed or incomplete^1,2^. Optimal management requires rapid recognition, timely initiation of intravenous corticosteroids (IVCS), structured monitoring, and early escalation to rescue therapy when indicated^3,4^. To standardize these high-stakes decisions, professional societies such as the American College of Gastroenterology (ACG) and the American Gastroenterological Association (AGA) have developed detailed clinical guidelines^3,5,6^. However, despite their availability, real-world adherence to ASUC guidelines remains variable^7,8^.

Monitoring guideline adherence, in general, is challenging. Current approaches rely on manual chart review which is time-consuming and expensive, or alternatively simple algorithms that analyze structured data alone, failing to realistically capture clinical complexity^9^. Deviations from recommended care are mostly detected retrospectively, limiting opportunities for timely intervention and quality improvement. Automated methods that can reliably apply guidelines to real-world clinical data could substantially improve real-time delivery of high-quality care while quantifying real-world guideline adherence^10^.

Recent studies are exploring the use of large language models (LLMs) for clinical decision support, with results showing that they can achieve high concordance with clinical guidelines or expert decision making^11–30^. However, important gaps remain. First, many published studies rely on synthetic vignettes rather than real electronic health records (EHR) data, making their results potentially overoptimistic^31^. Second, few investigations explicitly constrain models to predefined guidelines, leading to inconsistent or off-guideline outputs, potentially undermining clinical trust. Some studies have explored model adaptations like retrieval augmented generation (RAG)^17,20,28,32^ or custom GPT^19^ to enforce guideline adherence although off-guideline suggestions can still occur. Oniani et al^29^ and Deng et al^30^ both explored tighter guideline-constrained action space with binary tree traversal prompting but only reported performance on vignettes. Thirdly, automatic guideline application and adherence monitoring should be deterministic, transparent, and auditable along the cascade of decision making from recommendations to executions. Existing LLM-based approaches have not been systematically evaluated for this type of fine-grained, real-world guideline adherence monitoring of the cascade.

In this study, we address these three gaps by developing and validating an LLM framework for automated application and adherence monitoring of the 2025 ACG ASUC guidelines^3^ using real-world EHR data. We evaluate performance against manual chart review with a human performance ceiling defined by inter-rater reliability and apply the framework to characterize real-world adherence patterns. By focusing on a high-risk inpatient condition like ASUC and a clearly defined set of guideline items, this work aims to demonstrate how carefully designed LLM systems can enable scalable, reproducible, and clinically meaningful monitoring of guideline-based care.

## Materials and Methods

### Cohort Definition

The ASUC cohort at UCSF was identified using a structured EHR filter, namely, primary coded diagnosis of ulcerative colitis (UC) (K51) at a hospital admission (Figure 1). Patients were at least 18 years old at the time of the hospital admission and did not have a code for Crohn’s disease (K50) prior to the admission. This ensured that edge-cases with an unclear diagnosis were removed. We extracted the identified clinical notes as well as relevant structured data elements, such as laboratory orders and results, and medication orders and administrations, documented during the hospital stay. Structured data was transformed into text format using simple regular expressions. Information was aggregated chronologically, such that daily updates for each hospitalization were obtained.

**Figure 1:**
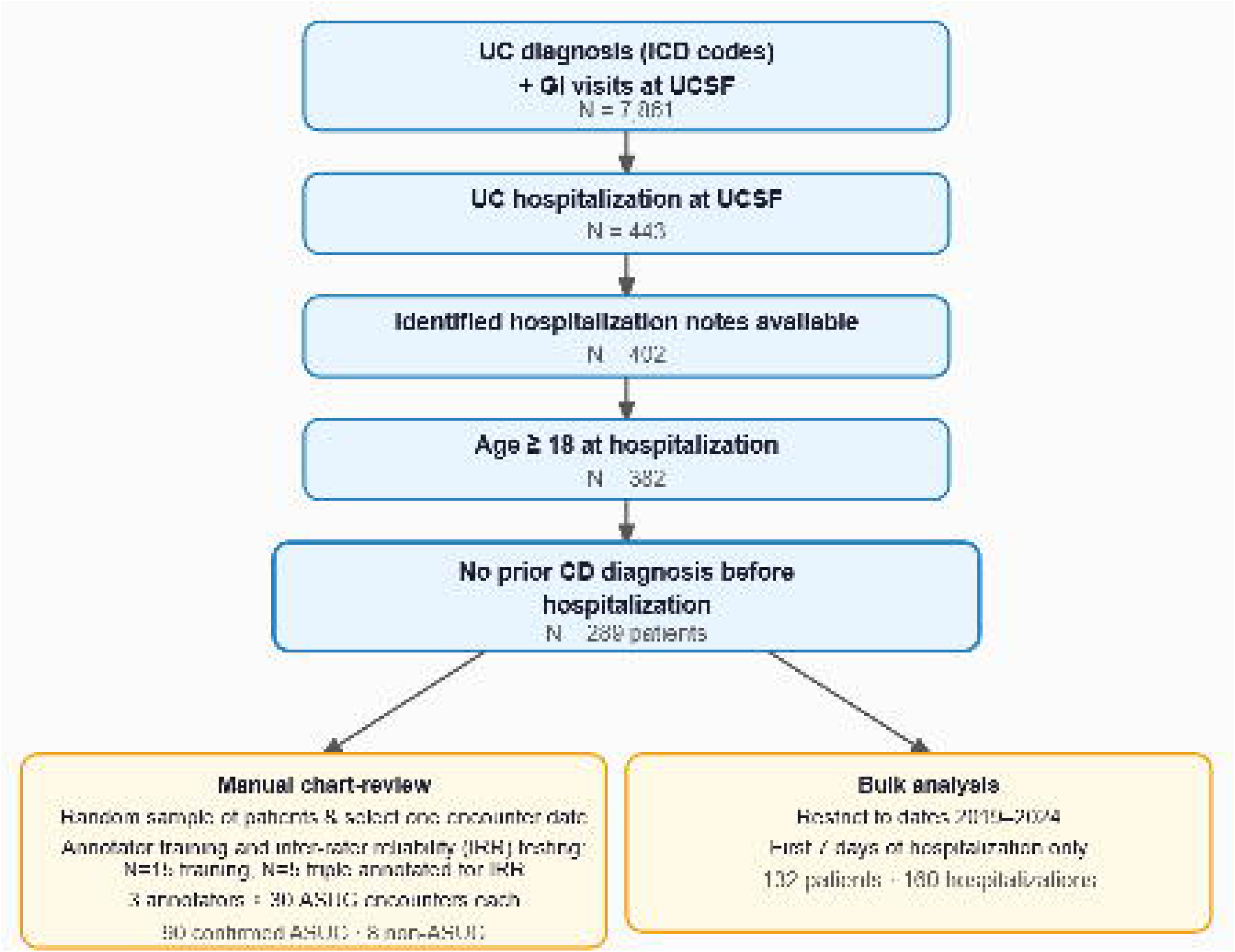
Patient Disposition Flowchart. The diagram depicts the decision process leading to our sample cohorts. From the identified ASUC hospitalization days we sampled one dataset for manual chart-review and validation of the LLM approach and one for a bulk real-world data analysis using the verified LLM approach. Abbreviations: ASUC, acute severe ulcerative colitis; CD: Crohn’s disease; GI, Gastroenterology; UC, ulcerative colitis Alt text: Flowchart showing how the cohort was selected to identify acute severe ulcerative colitis hospitalizations and how the sample cohort was split for manual chart review and validation.

### Manual Annotation

A labeled dataset was created through manual chart review of 103 randomly selected patients for a specific date during their hospital stays. Dates were selected to represent the whole hospital stay, from admission day until day of discharge. Manual annotation included an initial assessment of ASUC status and only proceeded if confirmed.

For relevant ASUC patient encounters on specific hospital days, we applied the 2025 ACG UC guidelines relevant to the management of ASUC, with nine relevant guideline items (recommendations 46–54, Table 1)^3^. For each ASUC guideline item, annotators identified binary status of whether it applied (A, predicates met), whether it was explicitly recommended by a gastroenterology consultant (R), whether guideline-compliant orders were placed (O), and whether the guideline-compliant care was executed (E). Three annotators underwent training and developed the annotation protocol (Supplemental Material) using 15 patient encounters not included in this analysis.

**Table 1:** Guideline-cascade agent performance on test set. The accuracy of the LLM versus single annotated manual chart review on the test set is depicted (N=60) for each guideline item and each variable (applies, recommended, ordered, executed). The downstream variables (recommended, ordered, executed) are only evaluated if ground truth applies = 1. We present the mean across five LLM calls and (min – max) to quantify reproducibility. Abbreviations: C. diff: *Clostridioides difficile*; DVT: deep vein thrombosis; IFX: infliximab; IVCS: intravenous corticosteroids; TPN: total parenteral nutrition

| Guideline Item | N (total /applies) | Applies Agreement % | Recommended Agreement % | Ordered Agreement % | Executed Agreement % |
|---|---|---|---|---|---|
| R46: C. diff testing | 60 / 60 | 99 (98–100) | 95 (93–95) | 95 (93–95) | 91 (88–92) |
| R47: DVT prophylaxis | 60 / 60 | 99 (98–100) | 91 (90–92) | 93 (92–93) | 85 (83–85) |
| R48: Avoid antibiotics | 60 / 60 | 99 (98–100) | 86 (85–87) | 96 (93–97) | 96 (95–97) |
| R49: Avoid TPN | 60 / 60 | 99 (98–100) | 96 (95–97) | 96 (95–97) | 99 (98–100) |
| R50: IVCS | 60 / 60 | 99 (98–100) | 97 (95– 98) | 97 (95–98) | 95 (93–97) |
| R51: Rescue therapy day 3 | 60 / 20 | 86 (83– 88) | 94 (90– 95) | 80 (80–80) | 80 (80–80) |
| R52: Maintain: IFX | 60 / 1 | 97 (97–100) | NA | NA | NA |
| R53: Maintain: thiopurines | 60 / 0 | 100 (100–100) | NA | NA | NA |
| R54: Maintain: vedolizumab | 60 / 0 | 100 (100–100) | NA | NA | NA |
| Overall (pooled) | 540 / 321 | 98 (97–99) | 93 (93–93) | 94 (93–95) | 92 (92–93) |

After training, inter-rater reliability was assessed on five example hospital days from five patients using Krippendorff’s alpha, prevalence, and raw agreement. The human performance ceiling was determined by first identifying the majority vote for each variable and subsequently evaluating each annotator against this gold standard. The mean accuracy of the three annotators against the majority vote served as the human performance ceiling. Across all evaluations only items where criteria A was true were evaluated for the downstream variables of recommendation, ordering, and execution. Each annotator reviewed and annotated 30 unique ASUC hospital days randomly sampled from multi-day hospitalization encounters. This process led to a total of 98 annotated hospital days (N=8 non-ASUC, N=90 ASUC).

### LLM Framework

We developed a multi-agent LLM framework to automate the application and assessment of adherence to the ACG guidelines relevant to the management of ASUC^3^. The workflow consisted of two primary agents: (1) a pre-selection agent that confirmed ASUC status, and (2) a guideline-cascade agent that, for each guideline item on a given hospital day, determined whether it applied and whether it was recommended by the clinician, ordered, and executed. This design enabled deterministic and auditable evaluation of predefined guidelines across the clinical workflow.

The LLM prompts were directly derived from the human annotation protocol to ensure consistency with expert evaluation (Supplemental Material). LLM inputs included the concatenated clinical notes from the hospital stay with injected text created from structured lab test order and results and medication data collected during the hospital visit. The input for a specific day only included data collected up until the end of that day. Two agents were designed, the pre-selection and the guideline-cascade agent, both using GPT-5 (version: 2025-08-07, knowledge cutoff: October 2024). LLMs were accessed via a HIPAA-compliant Azure API endpoint. Prompt engineering on a training set (N=30, 10 from each annotator) was performed with iterative improvements informed by error inspection.

Performance was evaluated on a held-out test set (N=60, 20 from each annotator). Metrics included accuracy, F1 score, sensitivity, and specificity. Due to varying prevalence, we report agreement percent as the main statistic. We performed five LLM runs with the same specifications to quantify reproducibility. Performance on the triple annotated data (N=5) was calculated against the majority vote and subsequently compared to the human performance ceiling. We assessed overall and item-specific performance as well as variable category (applies, recommended, ordered, executed) performance.

### Bulk Analysis for Real-World Adherence Monitoring

We evaluated real-world guideline adherence for the first seven days of all ASUC hospital stays documented between 2019 and 2024 using the validated prompts. First, the pre-selection agent confirmed ASUC status for each hospitalization based on the first available day. Those hospital stays that were confirmed to be ASUC were then processed with the guideline-cascade agent.

The agent identified the applicability of a given guideline item with all the available clinical information and annotated the full cascade from recommendation (R) to ordering (O) and execution (E). We investigated the workflow cascade, R → O → E, to identify where adherence stopped using the shorthand R±O±E±. We focused on four meaningful gap modes: failure to execute (R+O+E−), failure to order (R+O−E−), failure to recognize (R−O−E−), and silent adherence (R-O+E+). We estimated the conditional probability for proceeding with each step (e.g. P(O|R) as order follow-through given recommendation, P(E|O) as execution given order) and calculated confidence intervals with the Wilson method. We further computed the temporality of gaps by determining the number of gaps per hospital day (admission = day 1). We computed these for each cascade step (R, O, E) independently to identify patterns of workflow issues across the hospital stay.

We then focused on execution gaps, defined as instances where a guideline item applies, but the execution does not adhere to it regardless of recommendation and order status. We quantified these gaps and evaluated reasons for non-adherence. To identify the reason for execution gaps, clinical annotators defined categories for clinically justified (separate indication, contraindication, appropriate alternative, temporary hold) and clinically unjustified (patient refusal, process delay, simple violation) reasons and designed an LLM prompt (Supplemental Material). The input to the LLM consisted of the annotation instructions, the guideline where non-adherence was previously identified, and the original clinical documentation up to that hospital day. Structured output of the LLM included the identified category reason and free-text reason for non-adherence. This LLM prompt did not undergo systematic validation. We assessed whether reasons for non-adherence changed across the hospital stay by computing daily summaries.

### Ethics Declaration

This study was approved by the UCSF Institutional Review Board under Study #25-44405. All research was conducted in accordance with the U.S. Common Rule (45 CFR 46) and the principles of the Declaration of Helsinki. The UCSF IRB granted a waiver of informed consent for this retrospective study, as it posed no more than minimal risk and could not practicably be conducted without the waiver or without access to identifiable information.

## Results

### Annotation Reliability and Human Performance Ceiling

Three human annotators (AA, MYC, and SA) assessed all nine guideline items (recommendations 46-54, Supplementary Table 1) for each hospital day of the triple annotated data (N=5), overall demonstrating 93% raw agreement and 0.87 Krippendorff’s alpha (Supplementary Table 1). Near perfect agreement was demonstrated for indicating whether a guideline item applies (agreement = 0.96, alpha = 0.94). Alignment was lowest for whether the clinician explicitly recommended against routine antibiotics (R48, agreement = 0.6). Disagreements were generally only observed for guideline items of routine antibiotic avoidance (R48) and rescue therapy initiation (R51). Krippendorff’s alpha should be interpreted with caution given the class imbalance of some items and the low sample size. The human performance ceiling was derived by comparing each annotator against the majority vote (Supplementary Table 2). The mean accuracy across annotators against the majority vote was 0.97.

### Cohort Identification with ASUC Pre-selection Agent

After undergoing annotator training (N=15) and inter-rater reliability testing (N=5), the three human annotators reviewed 98 in-hospital days, sampled from a cohort identified with structured data filters, to confirm ASUC status and annotated for guideline applicability and adherence (Figure 1). Of these, 8 in-hospital days (8.2%) were adjudicated as non-ASUC by human reviewers.

Within this cohort pre-filtered by structured criteria for likely ASUC (N=98), the pre-selection agent achieved a sensitivity of 0.93 (false negatives = 6) and correctly rejected all 8 hospital days adjudicated as non-ASUC by manual chart-review. Because the evaluation cohort was enriched for ASUC and the non-ASUC hospital days were limited to structured-data false positives, specificity was not estimated from these data. Manual inspection of the false negatives revealed edge case scenarios where UC was considered as a contributor but not necessarily the main reason for the hospitalization when the patient was discharged, and later workup identified other main drivers for the symptoms. Overall, this indicates that the LLM can largely reproduce manual chart-review classification for cohort inclusion on top of structural data filters.

### LLMs Accurately Reproduce Expert Guideline Assessments

The guideline-cascade agent reached an average accuracy of 95% on the test set (Table 1, N=60) and matched the human-consensus performance ceiling on the triple annotated data (Supplemental Figure 1, Supplementary Table 2, N=5). On the test set, individual item-level performance was high across applicability (A), clinician recommendation (R), orders (O), and execution (E) for all nine guideline items, with applicability being the highest (98%). Lowest performance was observed for guideline R51 (rescue initiation, 85%) (Table 1) mirroring the lower human inter-rater agreement (Supplementary Table 1). Variability between LLM calls was minimal (Table 1).

### Real-World Guideline Adherence and Clinical Workflow Gap

Overall real-world adherence to ASUC guidelines was high. The first seven days of a total of 160 hospitalizations for 132 unique patients were assessed of which 121 hospitalizations for 104 patients were identified as ASUC by the pre-selection agent and thus 686 days were processed by the guideline-cascade agent. Guideline items R46–R50 commonly applied while item R51 applied in 40% of hospital days and R52–R54 never applied and were thus ignored for downstream analyses. This is because guideline items R52–R54 covers later stages of the hospitalization or outpatient maintenance care while we only examined adherence up until day 7 of the hospitalization.

Guideline non-adherence occurred across the clinical workflow. We leveraged our fine-grained annotation cascade that captured recommendation (R), order (O), and execution (E) to categorize eight possible gap modes that we denoted in shorthand as R±O±E± (Figure 2A, Supplementary Table 5) with R+O+E+ denoting full adherence. We further examined the transition probabilities from one step to the next (Supplementary Figure 2, Supplementary Table 6). Across applicable cases, full adherence (R+O+E+) was moderate to high for most guidelines (65–91%) except for R49 (0.1%).

**Figure 2:**
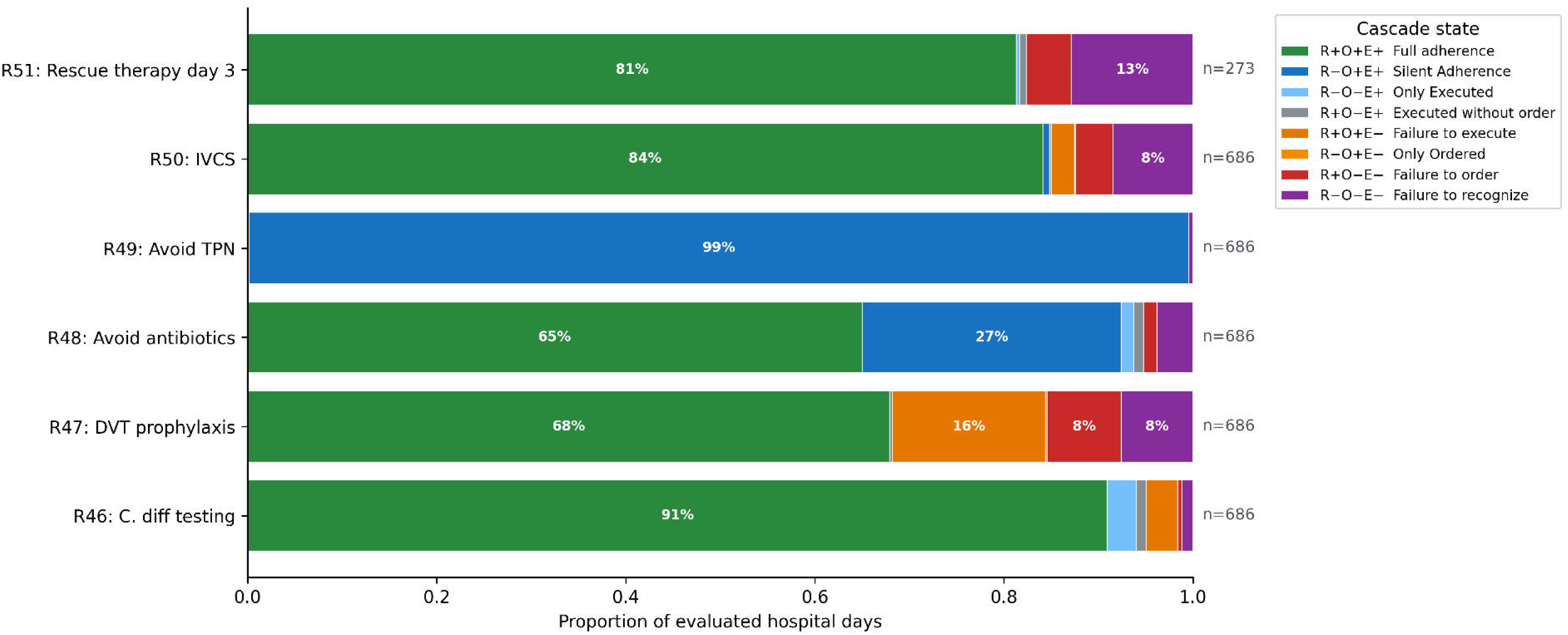
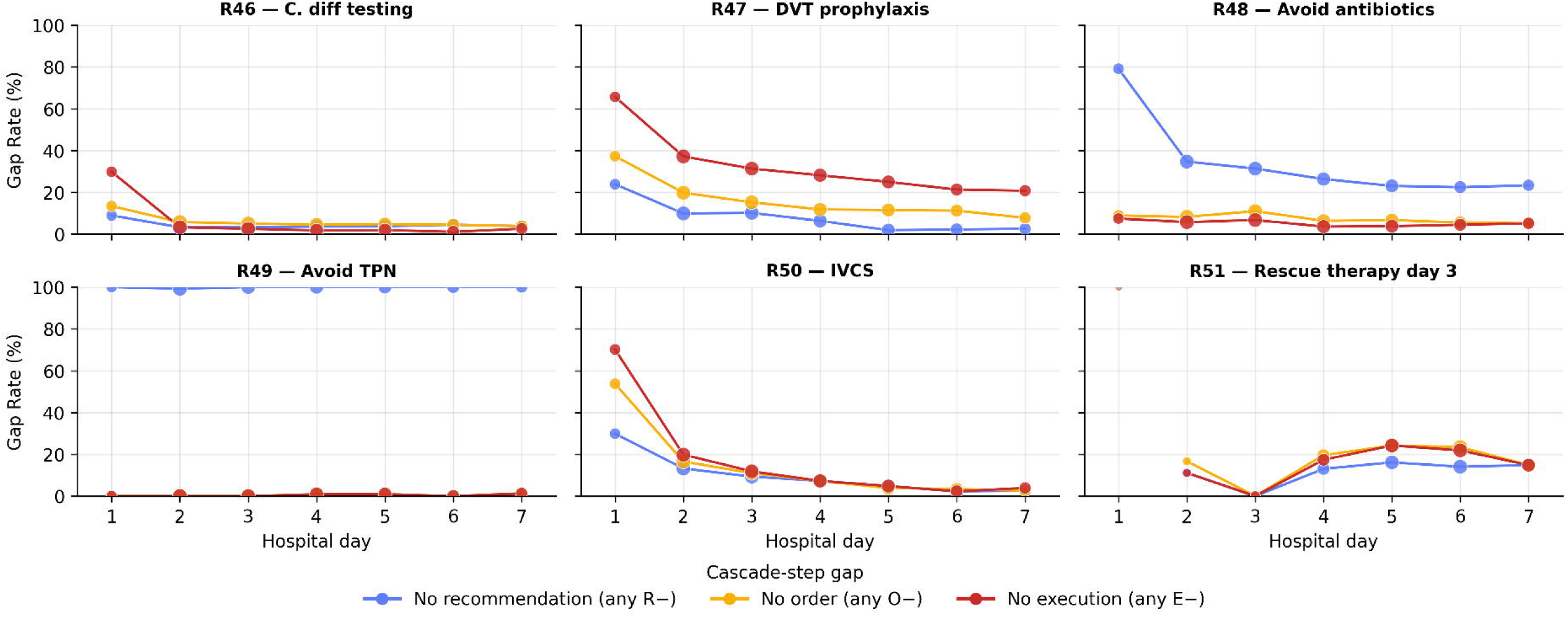
Gaps in Real-World Clinical Workflow. With the LLM agent, we examined in the real-world dataset where gaps in the clinical workflow occurred. Using a shorthand R±O±E± we investigated the proportion of each gap mode per guideline item (A) and when gaps occurred during the hospital stay (B). (A) The stacked bar plot shows the proportion of hospital days for which steps in the workflow (recommended, ordered, executed) followed the guidelines or not. (B) For each guideline the line chart shows the gap rate for each cascade step (R, O, E) for each hospital day. The grey numbers on the bottom display the number of applicable patients (denominator) according to the annotation of the LLM agents, which is also visualized by the size of the dots. annotation. Abbreviations: C. diff: *Clostridioides difficile*; DVT: deep vein thrombosis; IVCS: intravenous corticosteroids; TPN: total parenteral nutrition Alt text: Panel A is a stacked bar chart showing, for each guideline item, the proportion of hospital days in which recommendation, ordering and execution followed or deviated from the guideline. Panel B shows line charts of the daily gap rate for each step across the first seven hospital days.

The two avoidance guidelines (R48, R49) revealed a distinct adherence pattern driven by under-documentation rather than under-execution. For R49, TPN was almost always avoided in practice, but without any documented consultant deliberation, yielding a dominant R−O+E+ pattern (99% of applicable cases). R48 (avoid antibiotics) showed the same silent-adherence pattern less frequently (27% of hospital days), with the majority reaching full adherence. Recognition rates reflected this: high for "do" guidelines (87–96%) but substantially lower for avoidance items (68% for R48; 0.1% for R49). A key insight may thus be clinicians not documenting decisions to not act.

Downstream steps were highly reliable: ordering given recommendation was 91–100%, and execution given order was 96–100% for all but R47. R47, deep vein thrombosis (DVT) prophylaxis, was the only guideline for which “broke at execution” (R+O+E-) was the dominant gap mode (16%) reflecting missed execution despite appropriate recommendation and ordering. R50 (IVCS) and R51 (rescue therapy day 3) reached 84% and 81% full adherence respectively, with gaps concentrated at the recognition step (R−O−E−). A key vulnerability in ASUC care can thus be identified as inconsistent execution of DVT prophylaxis, with some gaps in timely initiation of IVCS and rescue therapy.

Temporal analysis of gap modes revealed three distinct failure-mode trajectories across guideline items (Figure 2B, Supplementary Table 7): early delays with recovery, persistent non-documentation, and conditional time-shifted patterns. The R46, R47, and R50 all showed deficits on the first days of the hospitalization but differed in how quickly and at which cascade step they resolved. R50 (IVCS) showed a pronounced day-1 deficit across recognition, ordering, and execution that resolved by day 3. Similarly, R46 (*C. diff* testing) showed a prominent day-1 execution gap (30%) despite much lower gap rates of recommendation and order, but it was nearly completely corrected by day 2. R47 non-adherence (DVT prophylaxis) followed a slower trajectory and only partial resolution, with execution gaps decreasing from 66% on day 1 to 21% by day 7. In contrast, avoidance guidelines showed higher recommendation gaps but steadily low order or execution gaps. The recommendation gap for R49 (TPN) remained consistently high throughout the hospital stay (>99%) due to non-documentation rather than delayed action, while R48 (antibiotics) exhibited persistent but gradually improving recognition gaps (day 1: 80%, day 7: 23%). Finally, R51 (rescue therapy) showed a time-shifted pattern reflecting its applicability after 72h of IV corticosteroid failure. Execution gaps were minimal during the first 48h post-admission (when the guideline was not yet clinically applicable in most cases), then rose to a peak of ∼24% around hospital days 4-5 before declining to ∼15% by day 7. This pattern suggests that gaps in rescue therapy delivery accumulate in the days following the 72h decision point.

### Reasons for Non-Adherence

Review of reasons for non-adherence, as classified by an LLM, showed that 62% of deviations were clinically unjustified, while the remainder reflected documented contraindications, separate therapeutic indications, or the use of appropriate alternative therapies (Figure 3). Justified deviations dominated the avoidance (R48, R49) and R51 guidelines. For R48 (avoid antibiotics), 64% of non-adherence reflected a separate therapeutic indication, consistent with antibiotics being appropriately prescribed for concurrent infections rather than inappropriately for ASUC. R49 (avoid TPN) showed only three gaps in total, all justified by separate therapeutic indication. For R51 (rescue therapy at day 3), 75% of non-adherence was due to the use of alternatives not included in the guideline as rescue therapies but reasonable in certain clinical scenarios, such as upadacitinib or other biologics in place of infliximab.

**Figure 3:**
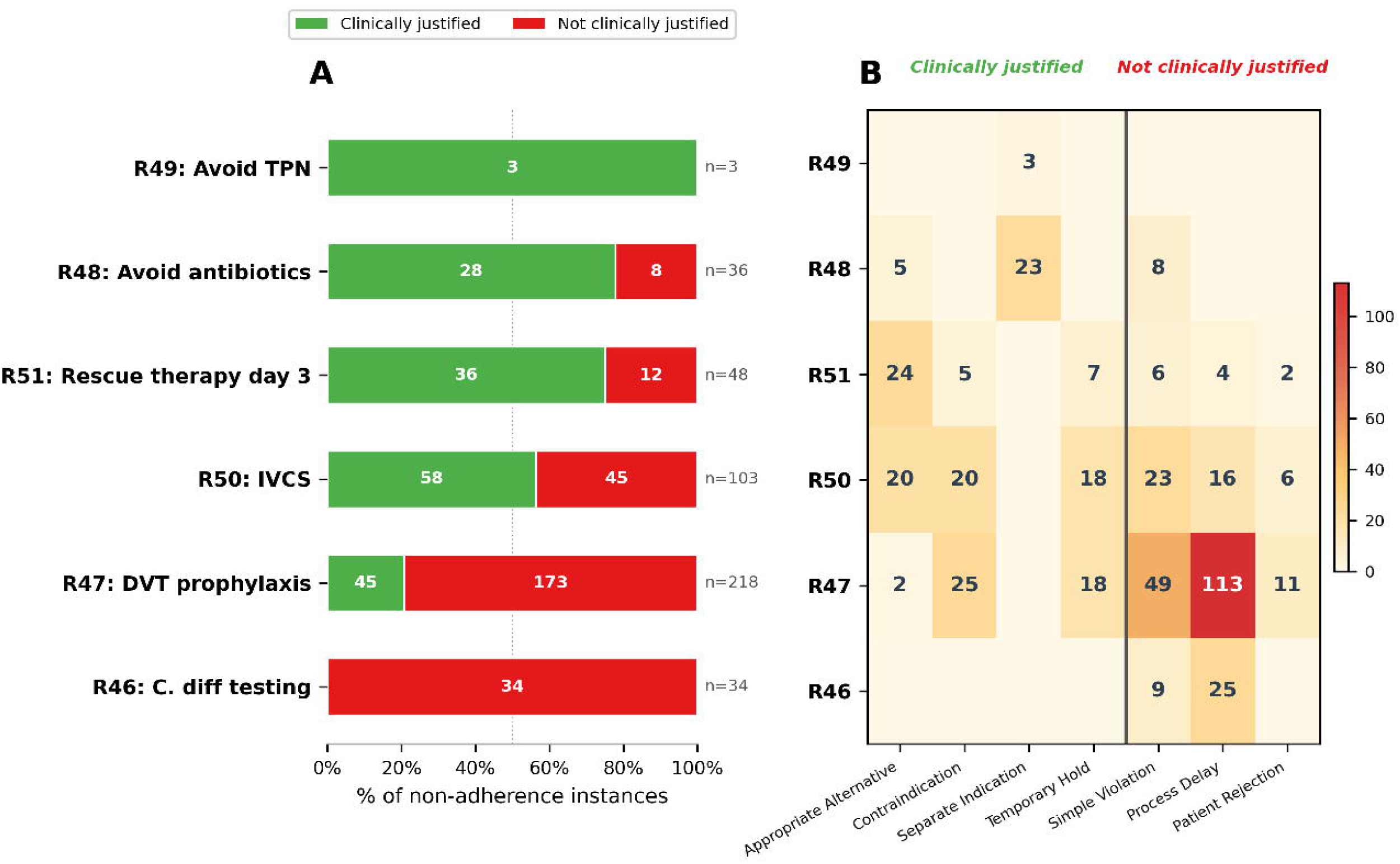
Reason for real-world non-adherence. On the real-world dataset N=444 execution gaps were identified and reanalyzed by an LLM to identify the reason for non-adherence. Seven categories were pre-defined. A) For each guideline item the total number of execution gaps is shown with the bar plot indicating the number of justified versus unjustified reasons. B) The heatmap depicts the number of violations per guideline item assigned to each reason category. Alt text: Panel A is a bar chart of execution gaps per guideline item, split into justified and unjustified deviations. Panel B is a heatmap of the number of deviations per guideline item across seven predefined reason categories.

On the other hand, clinically unjustified deviations concentrated in three guidelines: C. diff testing (100% of deviations unjustified), DVT prophylaxis (79%), and IVCS administration (44%) (Figure 3). DVT prophylaxis accounted for the largest absolute number of unjustified deviations (173 cases), most often reflecting process delays (65%). IVCS showed a more mixed profile across simple violations (51%), process delays (36%), and patient refusal, suggesting no single dominant failure mode. C. diff testing showed a similar pattern, with 74% of unjustified deviations attributable to process delays, likely reflecting early hospital days awaiting laboratory results.

The timing of reasons for non-adherence aligned with each guideline’s clinical action window (Supplementary Figure 3). For C. diff testing (R46), 59% of gaps occurred on hospital day 1 and were dominated by delays in placing an order. For rescue therapy (R51), the dominant reason across days 4–7 was appropriate alternative use, consistent with the guideline’s day-3 decision point and subsequent therapy choices. DVT prophylaxis (R47), which requires daily administration throughout the stay, showed gaps distributed across the hospital stay, with simple violations predominating in the first days and process delays thereafter. IVCS (R50) showed a temporal shift: day-1 gaps were disproportionately unjustified, while later gaps were enriched for clinically appropriate alternatives and contraindications. Notably, temporary holds clustered at the start of the stay, suggesting that IVCS initiation was sometimes deliberately delayed, likely due to pending C. diff results.

## Discussion

This study demonstrates that a multi-agent LLM framework can apply ASUC guidelines to real-world clinical scenarios at an expert level and resolve adherence into distinct workflow steps. The framework revealed documentation gaps for “avoid” guidelines and inconsistent execution of DVT prophylaxis as well as minor recognition gaps for IVCS and timely rescue therapy. Even though many deviations might be clinically justified, these patterns identify areas where decision support, reminders, or standardized workflows could further strengthen care delivery.

Our study is one of the first to examine LLM reasoning abilities at the intersection of practice guidelines and complex, real-world data. Most previous studies made use of artificial case vignettes to test LLM recommendations^13–15,17,19–27,29,33^, but a few have explored LLM treatment recommendations on real patient data^16,18,34,35^. On the other hand, most previous publications did not restrict the LLMs action space to the guidelines, while those that did specify the guidelines for the model did not use real-world data for evaluation^17,19,20,22,27,28^. This creates a unique gap that our work addresses.

Our real-world guideline adherence findings are consistent with prior work demonstrating gaps in adherence to established quality indicators in hospitalized ulcerative colitis. In a multicenter study, Nguyen et al.^40^ identified suboptimal adherence to key inpatient quality measures, including pharmacologic DVT prophylaxis, C. diff testing, and timely escalation of therapy, highlighting persistent variability in ASUC care. However, such previous studies were limited to aggregate assessments of whether recommended care was delivered, without the ability to localize where in the clinical workflow these gaps arose. In our study, we resolved adherence into its component steps and found multiple gaps and failure modes from recognition to execution. For example, we observed widespread "silent adherence" for avoidance-based recommendations: clinicians appeared to follow R48 (avoid routine antibiotics) and R49 (avoid TPN) in practice but rarely documented their deliberation. This likely reflects the implicit integration of these recommendations into routine care. On the other hand, the non-adherence to R50 (IVCS) and R51 (rescue therapy with infliximab) were concentrated at the recognition step and mostly related to the use of other biologics, reflecting a shift of practice towards newer biologics that can substitute for infliximab and/or IVCS pretreatment given recent evidence. Together, these findings underscore the importance of distinguishing true lapses in care from contextually appropriate decisions. Nuances can be missed by traditional adherence audits that treat any deviation as a failure.

Ensuring the ability of LLMs to consistently give clinical recommendations based on clinical guidelines is an important step towards a physician-facing clinical decision support and monitoring tool without user prompts. Guidelines are commonly considered the foundation of clinical decisions because they are highly standardized, expert-discussed, evidence-based protocols of clinical practice. Although current state-of-the-art clinical LLMs can provide guideline-based recommendations via free text to a certain extent, they are not primed to separate “purely guideline-bound" practice from additional clinical or systems-based considerations, unless carefully designed. The distinction is important in building the traceability, interpretability, and reliability of clinical AI. Therefore, validating LLM agents in this regard remains an active area of research.

Previous work has explored several strategies to enforce adherence to guidelines, including providing the full guideline in the prompt^22^, retrieval augmented generation (RAG)^17,20^, and custom GPTs combining guidelines in context with few-shot prompting and feedback^19^, all of which improve performance over naive suggestions. More stringent approaches including recursive binary decision tree traversal prompting^30^ and encoding conditional statements as code or network structures^29^, avoid off-guideline recommendations entirely but require transforming guidelines into specialized representations. Our approach similarly restricts recommendations to guidelines by constraining the LLM output to structured selections from guideline items provided in the prompt but only requires the recommendations in plain-text form as found in the guidelines, avoiding the need for decision-tree transformation and making the strategy easily adaptable across clinical domains. This ensures an adaptable system that can quickly integrate guideline updates, clinical practice changes, and be applied across clinical domains.

It is still noteworthy that guidelines are never meant to be the one and only source of practice guidance for a given patient. Multimorbidity, polypharmacy, and prior treatment failure, to name a few, are common complex issues in daily clinical care but are only incompletely addressed in guidelines^43^. In fact, guidelines cannot exhaustively enumerate all clinically plausible exceptions, nor can they fully capture the dynamic, context-dependent decision-making required at the bedside^44^. In addition, guidelines also represent a snapshot in time and may not include emerging trial data, off-label use, or institutional standards that have already changed clinical behavior^45^. These observations have direct implications for LLM-based decision support. Despite this caveat, any differences between guidelines and LLM recommendations should be justified separately, as our tool demonstrates in the semi-automatic LLM root cause analysis. The guideline component of LLM tools should also be adaptable to accommodate the ever-changing guidelines, which our tool also supports by incorporating free text. Beyond constraining recommendations, our approach also enables fine-grained assessment of adherence across the clinical workflow. Non-LLM adherence monitoring systems either operate on rule-based, single-step matching of executed interventions against recommendations^38^ or rely on manual chart review of aggregate quality indicators,^39^ which is time-consuming and not scalable. On the other hand, existing LLM-based adherence evaluations^30,36,37^ have assessed LLM outputs against guidelines in isolation without connecting them to actual clinical actions. We extended this by annotating not only which guidelines should be followed for a given patient, but also whether the clinical team recommended them, placed corresponding orders, and executed those orders. This cascade decomposition allows us to identify where in the workflow adherence breaks down, rather than treating it as a single binary outcome.

Several limitations should be noted. First, the study was conducted at a single academic center with a modest-sized gold standard dataset, which may limit generalizability to other institutions or community settings. Second, while the inputs to LLMs were based on a combination of clinical notes and structured elements, like orders and test results, other inputs, such as multidisciplinary discussion and undocumented verbal communications, are also important considerations but poorly captured in EHR. Third, adherence was assessed in literal interpretations of the nine guideline entries themselves. Namely, outside of the guideline texts, the accuracy of LLM outputs relies heavily on LLM’s “understanding” of clinical entities based on trained background domain knowledge. In addition, descriptions of summaries and strengths of evidence and recommendations from other guideline items, such as colectomy, were not included in the prompt. However, it is acknowledged that those nuances are important in human rationales and decisions. Fourth, while data from 2019–2024 was used to measure real-world adherence, the selected guidelines were published in 2025. Lastly, the LLM identifying the reasons for non-adherence was not formally validated.

Future work is needed to validate this framework across multiple health systems, guideline versions, and diseases. While our work operated on retrospective data, a real-time integration into clinical care is possible. A prospective study to assess clinical impact of real-time clinical guidance adherence monitoring is thereby warranted. Further encoding of guideline strength or adaptation of broad guidelines to meet individual practice pattern, such as accounting for formulary availability, institutional policies, or order execution pipelines, could be explored.

Overall, this study provides evidence that guideline-constrained LLM systems can achieve expert-level performance in applying ASUC guidelines and uncover adherence patterns at scale. These results support a promising path toward automated, reproducible, and clinically meaningful quality monitoring, with the potential to improve consistency among practitioners and result in better outcomes for patients with ASUC.

## Author contributions

VAR and UM designed the study, secured funding, supervised the work, and revised the manuscript. AKS developed the LLM framework, analyzed the data, and drafted the manuscript. MYC and AA contributed to the revision of LLM framework. BS, MYC, AA and SA developed the annotation protocol, performed chart review, and reviewed the manuscript. SL reviewed the manuscript. All authors approved the final version.

## Data availability statement

Code will be publicly available; data can be made available following a DUA with UCSF.

## Acknowledgements

The authors acknowledge the UCSF AI Tiger Team, Academic Research Services, Research Information Technology, and the Chancellor’s Task Force for Generative AI for their software development, analytical, and technical support related to the use of Versa API gateway (the UCSF secure implementation of large language models and generative AI via API gateway). We further thank the Facility for Advanced Computing (FAC) at UCSF for access to computation and data storage resources.

## Abbreviations

ACG: American College of Gastroenterology
AGA: American Gastroenterological Association
ASUC: acute severe ulcerative colitis
C. diff: Clostridioides difficile
DVT: deep vein thrombosis
EHR: electronic health record
IFX: infliximab
IVCS: intravenous corticosteroids
LLM: large language model
RAG: retrieval augmented generation
TPN: total parenteral nutrition
UC: ulcerative colitis
UCSF: University of California, San Francisco

