## Supplemental materials for "Real-world adherence to guidelines in acute severe ulcerative colitis: automated assessment using large language models"

Supplements

### Prompts and Protocols

#### Human annotation protocol

This protocol tells human reviewers how to extract and evaluate acute severe ulcerative-colitis (ASUC) management information from an **inpatient** clinical note (or admission packet) using the 2025 ACG guidelines.

Review **what is documented in APEX** —you may open other encounters, medication orders, imaging, or lab systems. If something is not explicitly documented, assume it did **not** occur.

For each provided patient, complete an excel sheet to check which guidelines apply, if they were recommended, if they were ordered according to guidelines (may equal ordered placed or may equal no order placed), and if they were executed at a specific timepoint during that patient’s admission (by 11:59pm of the service date). We want to see compliance with guidelines at a specific moment regardless of the reasons (even if they make clinical sense, e.g. GI bleed) or prior compliance.

There is a template excel sheets that you should use. They are stored in *insert folder location here*.

**ASUC ACG Guideline**

For each of the listed recommendations indicate:

1) Does the patient have ASUC? If not, then none of the guidelines apply. If they have ASUC, then review the following for each of the guidelines listed in the table below.
2) Does it Apply?
   a. Whether they apply (i.e. meet the criteria and thus should be executed). Generally, the first part of the sentence. Note that these guidelines are not written in a perfect fashion, so some details are left out and assumed to be known by the clinicians. Use your expert judgement on what criteria are entailed/implied.
   b. Whether an item applies depends on the **“if” (or subtle “if” condition(s)) of the item, e.g. maintain the remission achieved by xxx**.
   c. If a specific guideline does not apply, **all the following fields (recommended, ordered, executed) are N/A**
   d. Specific to guidelines
      i. If the patient has ASUC, R46-R50 apply
      ii. If the patient has failed to respond to steroids for 3 days, R51 applies (service date specific)
      iii. If patient received rescue therapy according to R51, **and** achieved clinical remission, R52-54 may apply. This is unlikely to occur unless the patient has a prolonged hospital stay. (service date specific)
3) Was it Recommended?
   a. Whether they were recommended by the consultant (did the attending recommend certain guidelines in their note)? This is commonly written in the assessment and plan section of the clinical note.
   b. Guideline needs to be specifically/explicitly stated. If consultant does not say anything, then they did not adhere to the guideline (then mark as ‘0’) (this goes for “avoid” statements as well) avoid statements include R48 and R49.
4) Was the order placed/not placed according to the guidelines? (Medication order, Lab Test Order, Diet Order)
   a. For certain guidelines (R47, R48) if there is no relevant order/ administration **COVERING** that exact calendar date (whether this is "remedied" within 24 hours or not, and whether there is an order/administration in the previous day), annotate this as **NOT** ordered/administered.
      i. R46 – At least one lab test must be ordered during admission (can be before service date)
      ii. R47 – Order must cover service date
      iii. R48 – Avoid statement, no order should be placed on service date
      iv. R49 – Avoid statement, no order should be placed on service date
      v. R50 – Order must cover service date
      vi. R51 – Order must be placed after 3 days of steroid use and before/on service date
5) Were the guidelines executed correctly?
   a. Placing an order does NOT equate to the guideline being executed (you need to see if there is documentation of the test/treatment being performed/Administered).
   b. Order should be placed before 23:59 of that calendar date to count
   c. If it is a lab test, see if results are available.
   d. If it pertains to medication, see if there is documentation that the medication was administered
      i. Please check the meds tab during the hospitalization and ensure the medications are administered using the “Administration” section.
      ii. For specific dosages that are mentioned, still denote them as “executed” even if the dosage/frequency does not perfectly align but state so in the “reason” column. In the ‘reason’ column, note down where you found the information in the chart or why you made that annotation. Consider all treatments/exams etc. performed during this hospitalization.
   e. For “avoid” statements indicate as 0 when they have been wrongly performed; in other words, indicate as 1 if they were AVOIDED CORRECTLY.
   f. For maintenance treatment (R52-54) also consider planned treatments denoted in A&P section.
      i. In some patients, they might be hospitalized long enough to make the items apply. Be sure to fill the “executed/ordered” columns in these patients.
   g. “Administration” fact in APeX is a starting point to look for the exact execution of inpatient orders
6) Reason columns
   a. For recommended, ordered and executed, in the associated ‘reason’ column, note down where you found the information in the chart or why you made that annotation.
      i. Recommended
         1. If 1: Date of note and line of text
         2. If 0: ‘Not recommended’
      ii. Ordered
          1. If 1: Date ordered and order number or for avoid guidelines, ‘not ordered’
          2. If 0: Reverse of 1
      iii. Executed
           1. If 1: Date executed and result (lab) or dosage (medication) or for avoid guidelines, ‘not executed’
           2. If 0: Reverse of 1

Multiple recommendations can apply and multiple can be executed, they’re not mutually exclusive.

| **ID** | **Exact wording** | **Cheat Sheet** |
| --- | --- | --- |
| R46 | In patients with ASUC, we recommend testing for C. difficile infection (Strong recommendation, moderate quality of evidence) | Order C. difficile PCR/EIA on admission.  Note: Must be performed during hospital admission. Make sure the **"results" in the left sidebar** or the **"results tab"** is double-checked. |
| R47 | In patients with ASUC, we recommend pharmacologic DVT prophylaxis as compared with no prophylaxis to prevent VTE (Strong recommendation, low quality of evidence) | Start LMWH (e.g., enoxaparin 40 mg SC daily, heparin) unless contraindicated.  Note: must be given on service date. |
| R48 | We recommend against routine use of broad-spectrum antibiotics in the management of ASUC (Strong recommendation, low quality of evidence). | Broad-spectrum antibiotics should not be given **specifically** for management of ASUC – may be provided for management of other concurrent conditions (ex: UTI, infection, etc.)  Note: avoid statement |
| R49 | We suggest against total parenteral nutrition for the purpose of bowel rest in ASUC (Conditional recommendation, very low quality of evidence) | Do not start TPN solely to “rest” the colon. “Solely” is an important consideration since some patients may receive TPN due to other reasons (should be elaborated on in the notes)  Note: avoid statement |
| R50 | In patients with ASUC, we recommend a total of 60 mg/d of methylprednisolone or hydrocortisone 100 mg 3 or 4 times per day to induce remission (Strong recommendation, low quality of evidence) | Begin high-dose IV steroids on day 1; note dosage if different (e.g., hydrocortisone 100 mg q6h–q8h or methylprednisolone 60 mg/d).  NOTE: please indicate discrepancies in dosage in the Reason column |
| R51 | In patients with ASUC failing to adequately respond to IVCS by 3 days, we recommend medical rescue therapy with infliximab or cyclosporine (Strong recommendation, moderate quality of evidence) | If no IVCS response by day 3, start infliximab 5 mg/kg or cyclosporine 2 mg/kg/d.  NOTE: Should receive infliximab after at least 72 hours of IVCS (no sooner). Guideline does not specify WHEN infliximab should be administered.  Need to complete at LEAST 3 days of steroids. If they were given with infliximab or cyclosporine earlier, then it is not in compliance. |
| R52 | In patients with ASUC who achieve remission with infliximab treatment, we recommend maintenance of remission with the same agent (Strong recommendations, moderate quality of evidence) | Continue scheduled infliximab (week 6, then q8 weeks). |
| R53 | In patients with ASUC who achieve remission with cyclosporine treatment, we suggest maintenance of remission with thiopurines (Conditional recommendation, low quality of evidence) | Start azathioprine or 6-MP once stable after induction. |
| R54 | In patients with ASUC who achieve remission with cyclosporine treatment, we suggest maintenance of remission with vedolizumab (Conditional recommendation, very low quality of evidence) | Switch to vedolizumab for maintenance once in remission. |

**Cheat-sheet for reviewers:**

- **IVCS failure clock** starts when IV steroid first given.
- **Rescue dosing**: infliximab 5 mg/kg; cyclosporine 2 mg/kg/d.
- **DVT prophylaxis**: e.g., enoxaparin 40 mg SC daily unless contraindicated.
  - Other terms to look at:
    - Heparin (not for flushing, should be standing or multiple doses), NOACs (rivaroxaban (Xarelto), apixaban (Eliquis), dabigatran (Pradaxa), edoxaban (Savaysa), or other"-xa" drugs).
- **CDI test** = PCR or EIA within admission
- Broad-spectrum abx count if > 24 h without documented infection.

Note: There appears to be usage of a template for guidelines/treatments at the end of the visit note (it appears in red). Even though it may be a template (not typed free-text), we will still consider this as recommended by the attending physician (hard to discern what was free text vs prefilled).

Edge Case:

- If a guideline was recommended by the consultant (in a conditional/futuristic tone), although the predicates were not yet satisfied up to that date, annotate as follows:
  - [0,1,0,0] (not applies, but recommended, not ordered, not executed)
- For any negative worded Guidelines (Ex: Guideline R48’s recommendation to AVOID broad-spectrum antibiotics solely for IBD and Guideline R49’s indicating to AVOID TPN), annotate as follows:
  - Applies = 1 if criteria are met
  - Recommended = 1 if explicitly stated to avoid
  - Ordered = 1 if there are no orders that would contradict this guideline
  - Executed = 1 if there are no administrations that would contradict this guideline
- If the C. diff test shows a detected toxin gene but no detectable toxin protein, the result will be considered negative (although based on our discussions, the results of the C. diff test don’t affect the rest of the guideline rules).

#### Pre-selection prompt

You are “UC-Severity-Assessor”, a severity assessment agent. You are an expert in ulcerative colitis (UC) and know all of the disease scoring systems like Truelove & Witt or the Mayo score. You can determine disease severity based on clinical notes.

Follow these structured steps:

- Identify the indication for the current hospitalization, summarize the full clinical course during this admission (including initial presentation, treatments given, and response), and determine both: the worst UC-related state during this hospitalization, and the most recent state at the time of the note.

- Reason about UC disease severity based primarily on the worst UC-related state during this hospitalization, while also taking into account the clinician’s judgement, reported symptoms, and lab tests.

- Determine whether the patient has acute severe ulcerative colitis (ASUC) during this hospitalization: Set ASUC = 1 if: the patient is hospitalized (in-patient status), AND a UC flare is the primary reason for the hospitalization (or clearly a major driver of admission). Set ASUC = 0 if: another diagnosis clearly explains the main symptoms and course (e.g., confirmed C. difficile infection, ischemic colitis, other primary non-UC process), even if the patient has a known UC diagnosis or ASUC is initially suspected, OR UC is clearly not the main driver of the current clinical problem.

Important rules for ASUC assignment:

- If the patient is admitted with a UC flare consistent with ASUC and then improves with treatment during the same hospitalization, you must still classify this admission as ASUC (ASUC = 1), as long as: the initial presentation met criteria for ASUC, AND no alternative primary diagnosis later clearly explains the symptoms better than UC.

- If the initial suspicion is ASUC but later in the hospitalization a definitive alternative primary diagnosis is reached (e.g., C. diff positive with symptoms attributed primarily to C. diff, or another colitis/infection judged to be the main cause), then set ASUC = 0 and clearly describe this reasoning in ASUC_reason.

Respond with only the JSON schema below, no extra text.

{

"disease_severity": "<not UC/Mild/Moderate/Moderate to Severe/Acute Severe/Unknown>",

"ASUC": "<0/1>",

"ASUC_reason": "<string>",

"stool_frequency":"<integer or string>",

"rectal_bleeding":"<string>",

"mucosal_appearance_endoscopy":"<string>",

"physician_rating":"<string>",

"temperature":"<float or string>",

"pulse":"<float or string>",

"hemoglobin":"<string>",

"Erythrocyte_sedimentation_rate":"<float or string>",

"additional_criteria_reasoning":["<list additional criteria met from notes>"]

}

#### Guideline-cascade prompt

This protocol explains how to extract and evaluate acute severe ulcerative-colitis (ASUC) management information from inpatient clinical notes and structured data extractions using the 2025 ACG guidelines. If something is not explicitly documented, assume it did not occur. For each provided patient at the specified timepoint, check which guidelines apply, if they were recommended, if they were ordered according to guidelines (may equal ordered placed or may equal no order placed), and if they were executed at a specific timepoint during that patient’s admission (by 11:59pm of the last service date). We want to see compliance with guidelines at a specific moment regardless of the reasons (even if they make clinical sense, e.g. GI bleed) or prior compliance.

**ASUC ACG Guideline**

R46. In patients with ASUC, we recommend testing for C. difficile infection

R47. In patients with ASUC, we recommend pharmacologic DVT prophylaxis as compared with no prophylaxis to prevent VTE

R48. We recommend against routine use of broad-spectrum antibiotics in the management of ASUC

R49. We suggest against total parenteral nutrition for the purpose of bowel rest in ASUC

R50. In patients with ASUC, we recommend a total of 60 mg/d of methylprednisolone or hydrocortisone 100 mg 3 or 4 times per day to induce remission

R51. In patients with ASUC failing to adequately respond to IVCS by 3 d, we recommend medical rescue therapy with infliximab or cyclosporine

R52. In patients with ASUC who achieve remission with infliximab treatment, we recommend maintenance of remission with the same agent

R53. In patients with ASUC who achieve remission with cyclosporine treatment, we suggest maintenance of remission with thiopurines

R54. In patients with ASUC who achieve remission with cyclosporine treatment, we suggest maintenance of remission with vedolizumab

For each of the listed recommendations indicate:

Does it Apply?

- Whether they apply (i.e. meet the criteria and thus should be executed). Generally, the first part of the sentence. Note that these guidelines are not written in a perfect fashion, so some details are left out and assumed to be known by the clinicians. Use your expert judgement on what criteria are entailed/implied.

- Whether an item applies depends on the “if” (or subtle “if” condition(s)) of the item, e.g. maintain the remission achieved by xxx.

- If a specific guideline does not apply, all the following fields (recommended, ordered, executed) are N/A

- Specific to guidelines

-- If the patient has ASUC, R46-R50 apply. Also suspected/probable ASUC counts or if UC flare was documented at any point duing admission.

-- If the patient has failed to respond to steroids for 3 consecutive days or rescue therapy is already initiated, R51 applies (service date specific)

-- If patient received rescue therapy according to R51, and achieved clinical remission, R52-54 may apply. This is unlikely to occur unless the patient has a prolonged hospital stay. (service date specific)

Was it Recommended?

- Whether they were recommended by the consultant (did the attending recommend certain guidelines in their note)? This is commonly written in the assessment and plan section of the clinical note.

- Guideline needs to be specifically/explicitly stated. If consultant does not say anything, then they did not adhere to the guideline (then mark as ‘0’) (this goes for “avoid” statements as well) avoid statements include R48 and R49.

- If orders were placed, consider this a recommendation (e.g. Cdiff testing ordered means it was also explicitly recommended).

Was the order placed/not placed according to the guidelines? (Medication order, Lab Test Order, Diet Order)

- For certain guidelines (R47, R48) if there is no relevant order/ administration COVERING that exact calendar date (whether this is "remedied" within 24 hours or not, and whether there is an order/administration in the previous day), annotate this as NOT ordered/administered.

- R46 – At least one lab test must be ordered during admission (can be before service date)

- R47 – Order must cover service date

- R48 – Avoid statement, no order should be placed covering service date without a documented non-IBD infectious indication

- R49 – Avoid statement, no order should be placed covering service date for bowel rest only

- R50 – Order must cover service date. Dosage can be ignored but must be IV steroid, not oral. If decided to be put on tapering schedule after positive response to IVCS or started on rescue therapy, then treat as =1.

- R51 – Order must cover the service date. For infliximab an order placed on a prior date counts as covering subsequent service dates during the same admission (no daily order required). For cyclosporine, the order must be active on the service date. Dosage can be ignored but must be intended for rescue therapy not reinitiation/continuation of usual medication.

Were the guidelines executed correctly?

- Placing an order does NOT equate to the guideline being executed (you need to see if there is documentation of the test/treatment being performed/Administered).

- Order should be placed before 23:59 of that calendar date to count

- for daily medications (e.g., IV steroids, DVT prophylaxis, cyclosporine): Executed = 1 only if administered on that service date. For steroids, if decided to put on taper after successful IVCS or started rescue, then treat as =1.

- for one-time or intermittent therapies (e.g., infliximab): Executed = 1 if administered on a prior service date of admission or on service date itself, subsequent service dates inherit executed=1.

- for Lab tests: Executed = 1 if result exists during admission, a lab result during the admission/immediately before counts as executed for the whole admission.

- Executed must be evaluated independently for each service date, exactly as ordered is. A missed administration on a prior date does not affect the executed annotation for the current service date.

- Please check the meds tab during the hospitalization and ensure the medications are administered using the “Administration” section.

- For specific dosages that are mentioned, still denote them as “executed” even if the dosage/frequency does not perfectly align but state so in the “reason” column. In the ‘reason’ column, note down where you found the information in the chart or why you made that annotation. Consider all treatments/exams etc. performed during this hospitalization. Do not confuse previous medications with current therapy and make sure intent is correct (ie infliximab used for rescue therapy).

- For “avoid” statements indicate as 0 when they have been wrongly performed; in other words, indicate as 1 if they were AVOIDED CORRECTLY.

- For maintenance treatment (R52-54) also consider planned treatments denoted in A&P section.

- In some patients, they might be hospitalized long enough to make the items apply. Be sure to fill the “executed/ordered” columns in these patients.

- “Administration” fact in APeX is a starting point to look for the exact execution of inpatient orders

Reason columns

For applies, recommended, ordered, and executed, in the associated ‘reason’ column, note down where you found the information in the chart or why you made that annotation.

- Applies

-- If 1: Date of note and line of text

-- If 0: Reverse of 1

- Recommended

-- If 1: Date of note and line of text

-- If 0: ‘Not recommended’

- Ordered

-- If 1: Date ordered and order number or for avoid guidelines, ‘not ordered’

-- If 0: Reverse of 1

- Executed

-- If 1: Date executed and result (lab) or dosage (medication) or for avoid guidelines, ‘not executed’

-- If 0: Reverse of 1

Multiple recommendations can apply and multiple can be executed, they’re not mutually exclusive.

Cheat-sheet for reviewers:

IVCS failure clock starts when IV steroid first given. Compliance with R51 is evaluated per service date. A patient is noncompliant on any service date ≥72 hours after IV steroid initiation if there is documented failure to respond and no rescue therapy has been initiated. Once rescue therapy is ordered/initiated, subsequent service dates are compliant.

Rescue dosing: infliximab 5 mg/kg; cyclosporine 2 mg/kg/d.

DVT prophylaxis: e.g., enoxaparin 40 mg SC daily unless contraindicated.

Other terms to look at:

Heparin (not for flushing, should be standing or multiple doses), NOACs (rivaroxaban (Xarelto), apixaban (Eliquis), dabigatran (Pradaxa), edoxaban (Savaysa), or other"-xa" drugs).

CDI test = PCR or EIA within admission; treat with oral vancomycin 125 mg q6h (See comment).

Broad-spectrum abx count if > 24 h without documented infection (Cdiff, UTI, etc).

Note: There appears to be usage of a template for guidelines/treatments at the end of the visit note (it appears in red). Even though it may be a template (not typed free-text), we will still consider this as recommended by the attending physician (hard to discern what was free text vs prefilled).

Edge Case:

If a guideline was recommended by the consultant (in a conditional/futuristic tone), although the predicates were not yet satisfied up to that date, annotate as follows:

[0,1,0,0] (not applies, but recommended, not ordered, not executed)

For any negative worded Guidelines (Ex: Guideline R48’s recommendation to AVOID broad-spectrum antibiotics solely for IBD and Guideline R49’s indicating to AVOID TPN), annotate as follows:

Applies = 1 if criteria are met

Recommended = 1 if explicitly stated to avoid

Ordered = 1 if there are no orders that would contradict this guideline

Executed = 1 if there are no administrations that would contradict this guideline

Output a structured JSON in this format:

{

"analysis": [

{

"guideline_id": "<string>",

"applies": <0 or 1>,

"applies_reasoning":"<string>",

"recommended": <0 or 1>,

"recommended_reasoning":"<string>",

"ordered": <0 or 1>,

"ordered_reasoning":"<string>",

"executed": <0 or 1>,

"executed_reasoning": "<string>"

},

...

],

}

"""

#### Reason for non-adherence prompt

You are an expert in identifying the reason for execution gaps in acute severe ulcerative colitis (ASUC) guideline adherence.

**Context**: You have already identified that a specific ACG guideline for ASUC management applied to this patient but was not executed by the service date. Your task is to determine WHY the guideline was not executed by examining clinical documentation. You will be given the guideline which was violated and the clinical documentation up until the service date as well as the reasoning from the previous assessment that identified an execution gap. The previous assessment's findings will be appended to the note under 'Previous assessment that identified this execution gap.' Use this as context.

**Execution Gap Categories** (select the most appropriate):

1. **Patient/Caregiver Rejection**

- Treatment was suggested/mentioned/discussed but patient/caregiver refused

2. **Contraindication**

- Clinical condition that makes the guideline dangerous/inappropriate/risky

- Includes documented relative contraindications when cited as the reason for non-execution

- Examples: severe thrombocytopenia or active bleeding (for DVT prophylaxis), documented severe allergy

- Use this when the contraindication is the PRIMARY reason — even if an alternative

was subsequently chosen as a result

3. **Temporary Hold**

- Explicitly documented as temporary

- Clear plan to resume (or resume implied by context, e.g., “held peri-op”)

- If the hold is due to a contraindication, prefer contraindication over temporary_hold

4. **Clinically Appropriate Alternative**

- Different agent or surgery used that satisfies guideline intent

- Clinically appropriate deviation from specific guideline recommendation

- Examples: UPA instead of IFX for rescue therapy, bowel resection/colectomy instead of further medicative treatment

- Use ONLY when the alternative was chosen on clinical/strategic grounds, NOT because the guideline-specified treatment was contraindicated or refused

5. **Process Delay**

- even though order in place, drug/test was not administered on the service date

- Could be a missed dose or a delayed start

- Use only when there is no documentation of a clinical decision not to execute the guideline; it is a process failure.

6. **Separate Therapeutic Indication**

- The intervention was initiated (or withheld) for a documented clinical indication distinct from the guideline’s intended therapeutic target.

- This includes treatment of concurrent conditions and treatment of ASUC-related complications (e.g., severe malnutrition) when the therapy is not being used to treat colitis itself

7. **Simple Violation of Guideline**

- Guideline clearly applicable with no documented contraindication or other reason

- Represents true non-adherence without clinical justification

- Examples: rescue therapy initiated on day 5 not day 3 without any justification

**JSON Output Format:**

{

"guideline_id": "<string>",

"service_date": "<date>",

"gap_reasons": {

"patient_caregiver_rejection": <0 or 1>,

"contraindication": <0 or 1>,

"temporary_hold": <0 or 1>,

"clinically_appropriate_alternative": <0 or 1>,

"process_delay": <0 or 1>,

"separate_therapeutic_indication": <0 or 1>,

"simple_violation": <0 or 1>

},

"primary_gap_reason": "patient_caregiver_rejection"|"contraindication"|"temporary_hold"|"clinically_appropriate_alternative"|"process_delay"|"separate_therapeutic_indication"|"simple_violation",

"clinical_justification": "<string: explanation of why guideline was not executed based on chart review>",

"supporting_evidence": [

{

"source": "<note type and date>",

"quote_or_finding": "<relevant text or observation>",

"timestamp": "<if available>"

}

]

}

**Important Notes:**

- Focus on what IS documented, not speculation about what might have happened

- "Simple Violation" should only be used when you've ruled out all other categories

- "primary_gap_reason" must be exactly one of the keys in "gap_reasons" set to 1.

- When determining the reason, evaluate documentation as of the service date. If a prior justification resolved before the service date and no new justification is documented, do not use that prior reason.

- A deviation from dosage is not a violation of guidelines. There may be another reason why non-compliance was annotated.

### Figures

#### Supplemental Figure 1: LLM reaches human-level performance

The gold-standard annotation on the triple annotated data (N=5) was determined as the majority vote across the three annotators. Mean accuracy of individual annotators (N=3) against the majority vote as well as LLM performance of all five runs against it was calculated. Only rows where gold standard annotation applies = 1 were considered for downstream variables (recommend, ordered, executed). The heatmap depicts the difference between human performance and LLM performance with <=0 indicating the LLM achieved human-level performance and >0 showing LLMs underperformed compared to humans. This is shown for each guideline item (R46-R54) and each variable (applies, recommended, ordered, executed) using mean human and mean LLM performance against the majority vote.

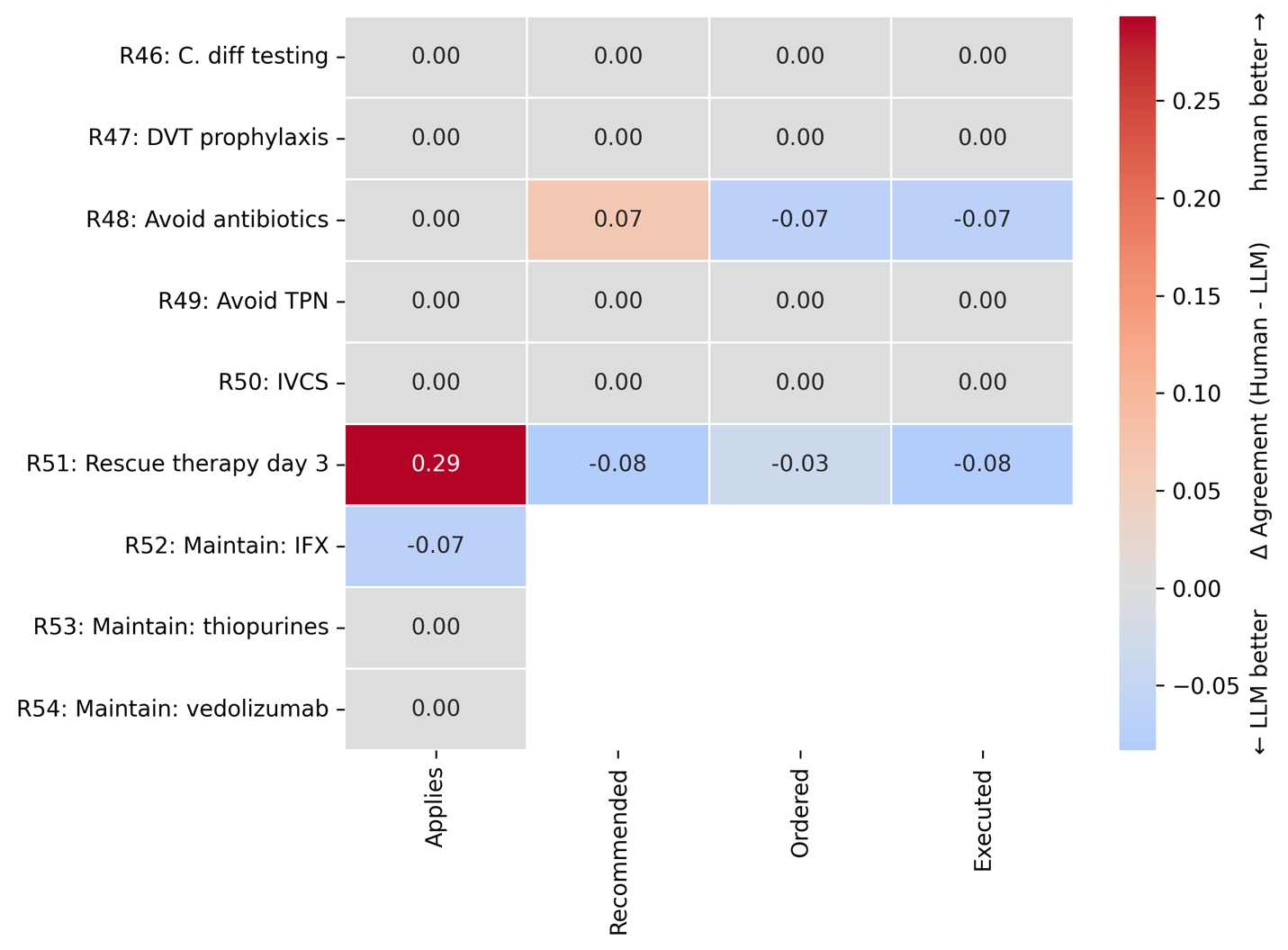

#### Supplemental Figure 2: Probability of conditional cascade steps

For each guideline the barplots indicate the probability of a next step in the workflow, meaning applies -> recommended -> ordered -> executed. The first step is recognizing that a guideline applies and recommending it accordingly (blue), then to follow-through with the order after that recommendation (yellow), and finally to execute the order (green). In grey we marked those that were executed even though the initial step of recommendation was missed.

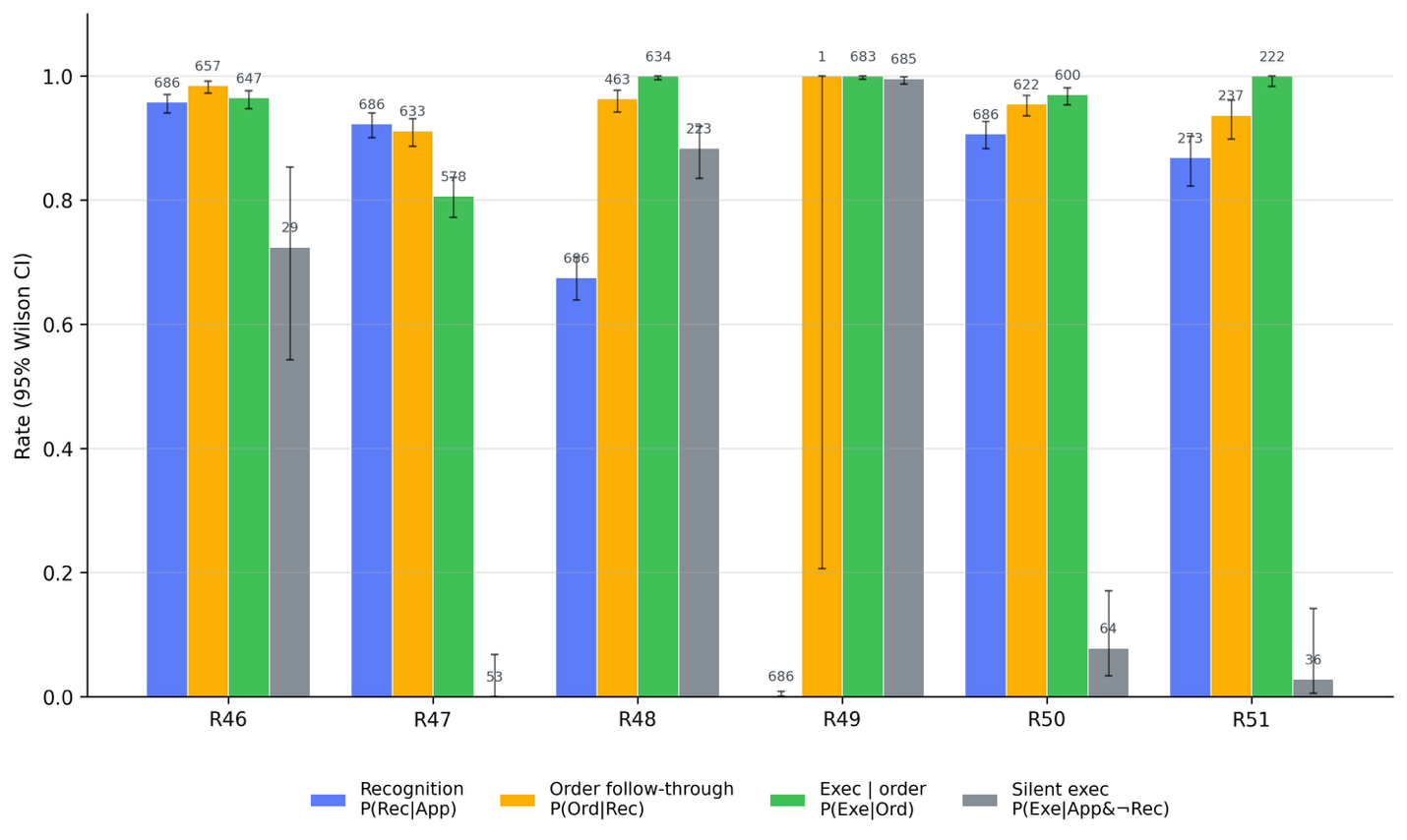

#### Supplementary Figure 3: Reasons for non-compliance changes over hospital stay

The stacked barplots show the number of execution gaps per hospital day indicating for the the proportion of reasons for the gap.

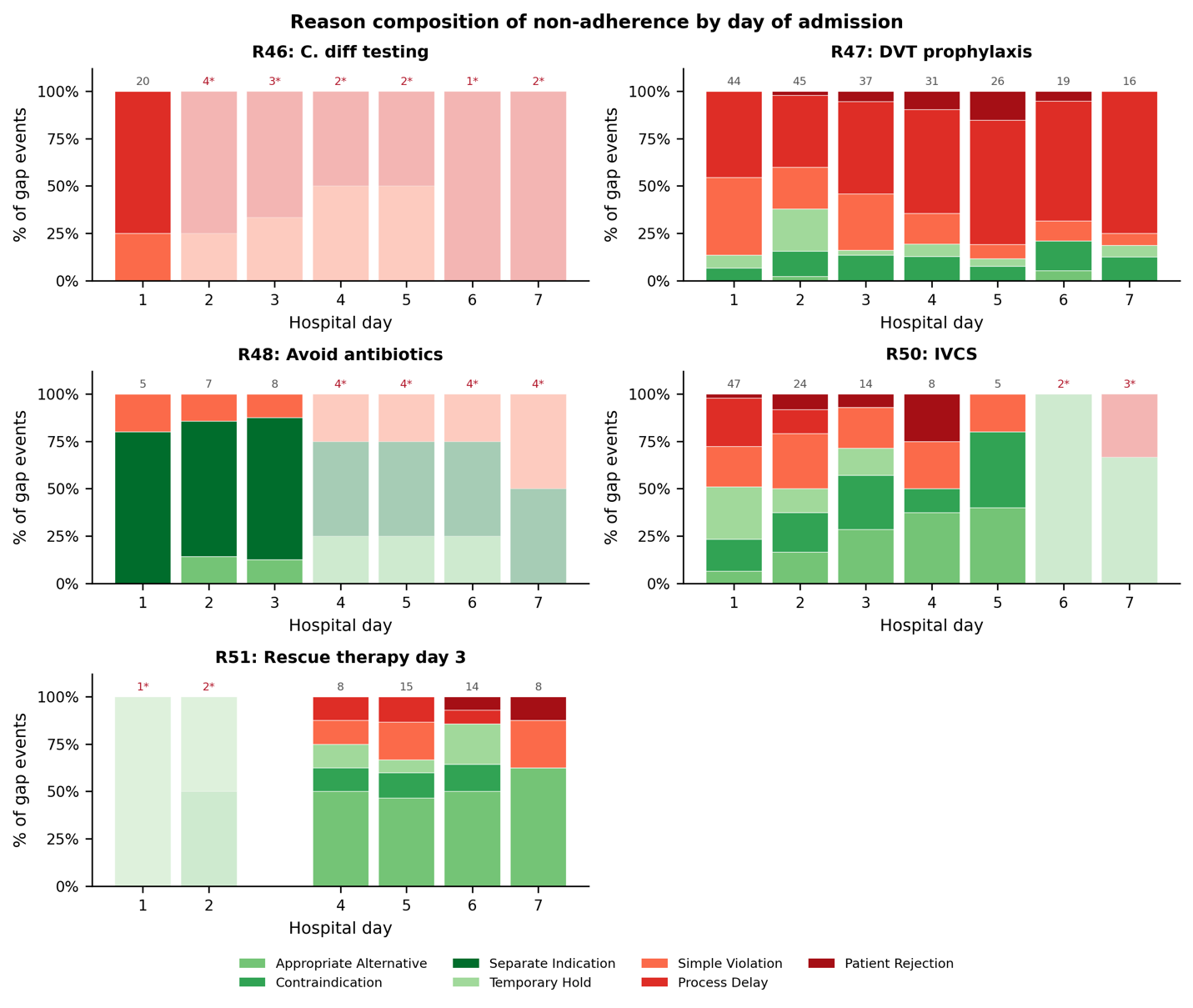

### Tables

#### Supplementary Table 1: Inter-annotator Agreement

Prevalence and raw agreement as well as Krippendorff’s alpha for each guideline item and variable as well as overall.

| variable | guideline | prevalence | agreement_rate | agrees | total | alpha |
| --- | --- | --- | --- | --- | --- | --- |
| Applies_yesno | R46 - Test for C. difficile infection | 1 | 1 | 5 | 5 | 1 |
| Applies_yesno | R47 - Provide pharmacologic DVT prophylaxis | 1 | 1 | 5 | 5 | 1 |
| Applies_yesno | R48 - Avoid routine broad-spectrum antibiotics | 1 | 1 | 5 | 5 | 1 |
| Applies_yesno | R49 - Avoid total parenteral nutrition solely ... | 1 | 1 | 5 | 5 | 1 |
| Applies_yesno | R50 - Induce remission with high-dose IV steroids | 1 | 1 | 5 | 5 | 1 |
| Applies_yesno | R51 - Rescue with infliximab or cyclosporine i... | 0.733333 | 0.8 | 4 | 5 | 0.681818 |
| Applies_yesno | R52 - Maintain remission achieved with inflixi... | 0.066667 | 0.8 | 4 | 5 | 0 |
| Applies_yesno | R53 - Maintain remission achieved with thiopur... | 0 | 1 | 5 | 5 | 1 |
| Applies_yesno | R54 - Maintain remission achieved with vedoliz... | 0 | 1 | 5 | 5 | 1 |
| Wasitrecommendedbytheconsultant_yesno | R46 - Test for C. difficile infection | 1 | 1 | 5 | 5 | 1 |
| Wasitrecommendedbytheconsultant_yesno | R47 - Provide pharmacologic DVT prophylaxis | 1 | 1 | 5 | 5 | 1 |
| Wasitrecommendedbytheconsultant_yesno | R48 - Avoid routine broad-spectrum antibiotics | 0.666667 | 0.6 | 3 | 5 | 0.44 |
| Wasitrecommendedbytheconsultant_yesno | R49 - Avoid total parenteral nutrition solely ... | 0 | 1 | 5 | 5 | 1 |
| Wasitrecommendedbytheconsultant_yesno | R50 - Induce remission with high-dose IV steroids | 1 | 1 | 5 | 5 | 1 |
| Wasitrecommendedbytheconsultant_yesno | R51 - Rescue with infliximab or cyclosporine i... | 0.666667 | 0.75 | 3 | 4 | 0.65625 |
| Orderplaced_yesno | R46 - Test for C. difficile infection | 1 | 1 | 5 | 5 | 1 |
| Orderplaced_yesno | R47 - Provide pharmacologic DVT prophylaxis | 1 | 1 | 5 | 5 | 1 |
| Orderplaced_yesno | R48 - Avoid routine broad-spectrum antibiotics | 0.933333 | 0.8 | 4 | 5 | 0 |
| Orderplaced_yesno | R49 - Avoid total parenteral nutrition solely ... | 1 | 1 | 5 | 5 | 1 |
| Orderplaced_yesno | R50 - Induce remission with high-dose IV steroids | 1 | 1 | 5 | 5 | 1 |
| Orderplaced_yesno | R51 - Rescue with infliximab or cyclosporine i... | 0.666667 | 0.75 | 3 | 4 | 0.65625 |
| Executed_yesno | R46 - Test for C. difficile infection | 1 | 1 | 5 | 5 | 1 |
| Executed_yesno | R47 - Provide pharmacologic DVT prophylaxis | 1 | 1 | 5 | 5 | 1 |
| Executed_yesno | R48 - Avoid routine broad-spectrum antibiotics | 0.933333 | 0.8 | 4 | 5 | 0 |
| Executed_yesno | R49 - Avoid total parenteral nutrition solely ... | 1 | 1 | 5 | 5 | 1 |
| Executed_yesno | R50 - Induce remission with high-dose IV steroids | 1 | 1 | 5 | 5 | 1 |
| Executed_yesno | R51 - Rescue with infliximab or cyclosporine i... | 0.666667 | 0.75 | 3 | 4 | 0.65625 |
| Applies_yesno | OVERALL | 0.644444 | 0.955556 | 43 | 45 | 0.935824 |
| Wasitrecommendedbytheconsultant_yesno | OVERALL | 0.724138 | 0.896552 | 26 | 29 | 0.829365 |
| Orderplaced_yesno | OVERALL | 0.942529 | 0.931034 | 27 | 29 | 0.580488 |
| Executed_yesno | OVERALL | 0.942529 | 0.931034 | 27 | 29 | 0.580488 |
| OVERALL | OVERALL | 0.792929 | 0.931818 | 123 | 132 | 0.861931 |

#### Supplementary Table 2: Human performance ceiling

Mean accuracy across annotators against the majority vote on the N=5 triple annotated data.

| Guideline Item | Applies | executed | Ordered | recommended |
| --- | --- | --- | --- | --- |
| R46 - Test for C. difficile infection | 1.000000 | 1.000000 | 1.000000 | 1.000000 |
| R47 - Provide pharmacologic DVT prophylaxis | 1.000000 | 1.000000 | 1.000000 | 1.000000 |
| R48 - Avoid routine broad-spectrum antibiotics | 1.000000 | 0.933333 | 0.933333 | 0.866667 |
| R49 - Avoid total parenteral nutrition solely for bowel rest | 1.000000 | 1.000000 | 1.000000 | 1.000000 |
| R50 - Induce remission with high-dose IV steroids | 1.000000 | 1.000000 | 1.000000 | 1.000000 |
| R51 - Rescue with infliximab or cyclosporine if no IVCS response by day 3 | 0.933333 | 0.916667 | 0.916667 | 0.916667 |
| R52 - Maintain remission achieved with infliximab, if infliximab used for rescue | 0.933333 | NaN | NaN | NaN |
| R53 - Maintain remission achieved with thiopurines, if cyclosporine used for rescue | 1.000000 | NaN | NaN | NaN |
| R54 - Maintain remission achieved with vedolizumab, if cyclosporine used for rescue | 1.000000 | NaN | NaN | NaN |

#### Supplementary Table 3: Performance on train set

| GuidelineID | Category | N | Prevalence | Pct_Agreement | F1 | Precision | Recall | TP | FP | FN | TN |
| --- | --- | --- | --- | --- | --- | --- | --- | --- | --- | --- | --- |
| R46: C. diff testing | applies | 30 | 1 | 1 | 1 | 1 | 1 | 30 | 0 | 0 | 0 |
| R46: C. diff testing | recommended | 30 | 1 | 1 | 1 | 1 | 1 | 30 | 0 | 0 | 0 |
| R46: C. diff testing | ordered | 30 | 0.967 | 0.967 | 0.983 | 0.967 | 1 | 29 | 1 | 0 | 0 |
| R46: C. diff testing | executed | 30 | 0.933 | 0.967 | 0.982 | 0.966 | 1 | 28 | 1 | 0 | 1 |
| R47: DVT prophylaxis | applies | 30 | 1 | 1 | 1 | 1 | 1 | 30 | 0 | 0 | 0 |
| R47: DVT prophylaxis | recommended | 30 | 0.9 | 0.933 | 0.964 | 0.931 | 1 | 27 | 2 | 0 | 1 |
| R47: DVT prophylaxis | ordered | 30 | 0.867 | 0.9 | 0.943 | 0.926 | 0.962 | 25 | 2 | 1 | 2 |
| R47: DVT prophylaxis | executed | 30 | 0.7 | 0.967 | 0.976 | 1 | 0.952 | 20 | 0 | 1 | 9 |
| R48: Avoid antibiotics | applies | 30 | 1 | 1 | 1 | 1 | 1 | 30 | 0 | 0 | 0 |
| R48: Avoid antibiotics | recommended | 30 | 0.533 | 0.833 | 0.857 | 0.789 | 0.938 | 15 | 4 | 1 | 10 |
| R48: Avoid antibiotics | ordered | 30 | 0.9 | 0.9 | 0.945 | 0.929 | 0.963 | 26 | 2 | 1 | 1 |
| R48: Avoid antibiotics | executed | 30 | 0.9 | 0.9 | 0.945 | 0.929 | 0.963 | 26 | 2 | 1 | 1 |
| R49: Avoid TPN | applies | 30 | 1 | 1 | 1 | 1 | 1 | 30 | 0 | 0 | 0 |
| R49: Avoid TPN | recommended | 30 | 0 | 1 | 0 | 0 | 0 | 0 | 0 | 0 | 30 |
| R49: Avoid TPN | ordered | 30 | 0.933 | 0.867 | 0.926 | 0.962 | 0.893 | 25 | 1 | 3 | 1 |
| R49: Avoid TPN | executed | 30 | 0.933 | 0.9 | 0.945 | 0.963 | 0.929 | 26 | 1 | 2 | 1 |
| R50: IV steroids | applies | 30 | 1 | 1 | 1 | 1 | 1 | 30 | 0 | 0 | 0 |
| R50: IV steroids | recommended | 30 | 0.967 | 0.933 | 0.964 | 1 | 0.931 | 27 | 0 | 2 | 1 |
| R50: IV steroids | ordered | 30 | 0.933 | 0.967 | 0.982 | 1 | 0.964 | 27 | 0 | 1 | 2 |
| R50: IV steroids | executed | 30 | 0.933 | 0.967 | 0.982 | 1 | 0.964 | 27 | 0 | 1 | 2 |
| R51: Rescue therapy day 3 | applies | 30 | 0.467 | 0.7 | 0.69 | 0.667 | 0.714 | 10 | 5 | 4 | 11 |
| R51: Rescue therapy day 3 | recommended | 14 | 0.5 | 0.929 | 0.923 | 1 | 0.857 | 6 | 0 | 1 | 7 |
| R51: Rescue therapy day 3 | ordered | 14 | 0.357 | 0.786 | 0.727 | 0.667 | 0.8 | 4 | 2 | 1 | 7 |
| R51: Rescue therapy day 3 | executed | 14 | 0.357 | 0.786 | 0.727 | 0.667 | 0.8 | 4 | 2 | 1 | 7 |
| R52: Maintain: IFX | applies | 30 | 0 | 1 | 0 | 0 | 0 | 0 | 0 | 0 | 30 |
| R52: Maintain: IFX | recommended | 0 |  |  |  |  |  | 0 | 0 | 0 | 0 |
| R52: Maintain: IFX | ordered | 0 |  |  |  |  |  | 0 | 0 | 0 | 0 |
| R52: Maintain: IFX | executed | 0 |  |  |  |  |  | 0 | 0 | 0 | 0 |
| R53: Maintain: thiopurines | applies | 30 | 0 | 1 | 0 | 0 | 0 | 0 | 0 | 0 | 30 |
| R53: Maintain: thiopurines | recommended | 0 |  |  |  |  |  | 0 | 0 | 0 | 0 |
| R53: Maintain: thiopurines | ordered | 0 |  |  |  |  |  | 0 | 0 | 0 | 0 |
| R53: Maintain: thiopurines | executed | 0 |  |  |  |  |  | 0 | 0 | 0 | 0 |
| R54: Maintain: vedolizumab | applies | 30 | 0 | 1 | 0 | 0 | 0 | 0 | 0 | 0 | 30 |
| R54: Maintain: vedolizumab | recommended | 0 |  |  |  |  |  | 0 | 0 | 0 | 0 |
| R54: Maintain: vedolizumab | ordered | 0 |  |  |  |  |  | 0 | 0 | 0 | 0 |
| R54: Maintain: vedolizumab | executed | 0 |  |  |  |  |  | 0 | 0 | 0 | 0 |
| Overall | applies | 270 | 0.607 | 0.967 | 0.973 | 0.97 | 0.976 | 160 | 5 | 4 | 101 |
| Overall | recommended | 164 | 0.665 | 0.939 | 0.955 | 0.946 | 0.963 | 105 | 6 | 4 | 49 |
| Overall | ordered | 164 | 0.872 | 0.909 | 0.948 | 0.944 | 0.951 | 136 | 8 | 7 | 13 |
| Overall | executed | 164 | 0.835 | 0.927 | 0.956 | 0.956 | 0.956 | 131 | 6 | 6 | 21 |
| Overall | all (pooled) | 762 | 0.726 | 0.94 | 0.959 | 0.955 | 0.962 | 532 | 25 | 21 | 184 |
| Overall | all (macro avg) | 762 | 0.745 | 0.936 | 0.958 | 0.954 | 0.962 | 532 | 25 | 21 | 184 |

#### Supplementary Table 4: Performance on test set

The TP, FP, FN, TN are the median values across the five runs. We depict the mean and the min, max values of the five LLM runs.

| GuidelineID | Category | N | Prevalence | Pct_Agreement | F1 | Precision | Recall | TP | FP | FN | TN |
| --- | --- | --- | --- | --- | --- | --- | --- | --- | --- | --- | --- |
| R46: C. diff testing | applies | 60 | 1 | 0.993 (0.983-1.000) | 0.997 (0.992-1.000) | 1.000 (1.000-1.000) | 0.993 (0.983-1.000) | 60 | 0 | 0 | 0 |
| R46: C. diff testing | recommended | 60 | 0.933 | 0.947 (0.933-0.950) | 0.972 (0.965-0.974) | 0.949 (0.948-0.949) | 0.996 (0.982-1.000) | 56 | 3 | 0 | 1 |
| R46: C. diff testing | ordered | 60 | 0.933 | 0.947 (0.933-0.950) | 0.972 (0.964-0.973) | 0.965 (0.964-0.965) | 0.979 (0.964-0.982) | 55 | 2 | 1 | 2 |
| R46: C. diff testing | executed | 60 | 0.917 | 0.910 (0.883-0.917) | 0.951 (0.937-0.955) | 0.943 (0.929-0.946) | 0.960 (0.945-0.964) | 53 | 3 | 2 | 2 |
| R47: DVT prophylaxis | applies | 60 | 1 | 0.993 (0.983-1.000) | 0.997 (0.992-1.000) | 1.000 (1.000-1.000) | 0.993 (0.983-1.000) | 60 | 0 | 0 | 0 |
| R47: DVT prophylaxis | recommended | 60 | 0.9 | 0.913 (0.900-0.917) | 0.953 (0.945-0.955) | 0.930 (0.929-0.930) | 0.978 (0.963-0.981) | 53 | 4 | 1 | 2 |
| R47: DVT prophylaxis | ordered | 60 | 0.8 | 0.930 (0.917-0.933) | 0.956 (0.947-0.958) | 0.962 (0.957-0.978) | 0.950 (0.938-0.958) | 46 | 2 | 2 | 10 |
| R47: DVT prophylaxis | executed | 60 | 0.7 | 0.847 (0.833-0.850) | 0.884 (0.875-0.886) | 0.941 (0.921-0.946) | 0.833 (0.833-0.833) | 35 | 2 | 7 | 16 |
| R48: Avoid antibiotics | applies | 60 | 1 | 0.993 (0.983-1.000) | 0.997 (0.992-1.000) | 1.000 (1.000-1.000) | 0.993 (0.983-1.000) | 60 | 0 | 0 | 0 |
| R48: Avoid antibiotics | recommended | 60 | 0.5 | 0.860 (0.850-0.867) | 0.865 (0.857-0.871) | 0.834 (0.818-0.844) | 0.900 (0.900-0.900) | 27 | 5 | 3 | 25 |
| R48: Avoid antibiotics | ordered | 60 | 0.967 | 0.957 (0.933-0.967) | 0.977 (0.965-0.983) | 0.983 (0.982-0.983) | 0.972 (0.948-0.983) | 57 | 1 | 1 | 1 |
| R48: Avoid antibiotics | executed | 60 | 0.967 | 0.960 (0.950-0.967) | 0.979 (0.974-0.983) | 0.983 (0.982-0.983) | 0.976 (0.966-0.983) | 57 | 1 | 1 | 1 |
| R49: Avoid TPN | applies | 60 | 1 | 0.993 (0.983-1.000) | 0.997 (0.992-1.000) | 1.000 (1.000-1.000) | 0.993 (0.983-1.000) | 60 | 0 | 0 | 0 |
| R49: Avoid TPN | recommended | 60 | 0.033 | 0.957 (0.950-0.967) | 0.000 (0.000-0.000) | 0.000 (0.000-0.000) | 0.000 (0.000-0.000) | 0 | 1 | 2 | 57 |
| R49: Avoid TPN | ordered | 60 | 0.983 | 0.957 (0.950-0.967) | 0.977 (0.974-0.983) | 1.000 (1.000-1.000) | 0.956 (0.949-0.966) | 56 | 0 | 3 | 1 |
| R49: Avoid TPN | executed | 60 | 0.983 | 0.987 (0.983-1.000) | 0.993 (0.991-1.000) | 1.000 (1.000-1.000) | 0.986 (0.983-1.000) | 58 | 0 | 1 | 1 |
| R50: IVCS | applies | 60 | 1 | 0.993 (0.983-1.000) | 0.997 (0.992-1.000) | 1.000 (1.000-1.000) | 0.993 (0.983-1.000) | 60 | 0 | 0 | 0 |
| R50: IVCS | recommended | 60 | 0.9 | 0.967 (0.950-0.983) | 0.982 (0.972-0.991) | 0.968 (0.964-0.982) | 0.996 (0.981-1.000) | 54 | 2 | 0 | 4 |
| R50: IVCS | ordered | 60 | 0.9 | 0.970 (0.950-0.983) | 0.984 (0.972-0.991) | 0.971 (0.964-0.982) | 0.996 (0.981-1.000) | 54 | 2 | 0 | 4 |
| R50: IVCS | executed | 60 | 0.9 | 0.953 (0.933-0.967) | 0.974 (0.963-0.981) | 0.971 (0.963-0.981) | 0.978 (0.963-0.981) | 53 | 2 | 1 | 4 |
| R51: Rescue therapy day 3 | applies | 60 | 0.333 | 0.863 (0.833-0.883) | 0.814 (0.762-0.844) | 0.744 (0.727-0.760) | 0.900 (0.800-0.950) | 18 | 6 | 2 | 34 |
| R51: Rescue therapy day 3 | recommended | 20 | 0.65 | 0.940 (0.900-0.950) | 0.955 (0.923-0.963) | 0.927 (0.923-0.929) | 0.985 (0.923-1.000) | 13 | 1 | 0 | 6 |
| R51: Rescue therapy day 3 | ordered | 20 | 0.45 | 0.800 (0.800-0.800) | 0.800 (0.800-0.800) | 0.727 (0.727-0.727) | 0.889 (0.889-0.889) | 8 | 3 | 1 | 8 |
| R51: Rescue therapy day 3 | executed | 20 | 0.45 | 0.800 (0.800-0.800) | 0.800 (0.800-0.800) | 0.727 (0.727-0.727) | 0.889 (0.889-0.889) | 8 | 3 | 1 | 8 |
| R52: Maintain: IFX | applies | 60 | 0.017 | 0.973 (0.967-1.000) | 0.600 (0.500-1.000) | 0.467 (0.333-1.000) | 1.000 (1.000-1.000) | 1 | 2 | 0 | 57 |
| R52: Maintain: IFX | recommended | 1 | 1 | NA | NA | NA | NA | 1 | 0 | 0 | 0 |
| R52: Maintain: IFX | ordered | 1 | 0 | NA | NA | NA | NA | 0 | 1 | 0 | 0 |
| R52: Maintain: IFX | executed | 1 | 0 | NA | NA | NA | NA | 0 | 0 | 0 | 1 |
| R53: Maintain: thiopurines | applies | 60 | 0 | 1.000 (1.000-1.000) | 0.000 (0.000-0.000) | 0.000 (0.000-0.000) | 0.000 (0.000-0.000) | 0 | 0 | 0 | 60 |
| R53: Maintain: thiopurines | recommended | 0 |  | NA | NA | NA | NA | 0 | 0 | 0 | 0 |
| R53: Maintain: thiopurines | ordered | 0 |  | NA | NA | NA | NA | 0 | 0 | 0 | 0 |
| R53: Maintain: thiopurines | executed | 0 |  | NA | NA | NA | NA | 0 | 0 | 0 | 0 |
| R54: Maintain: vedolizumab | applies | 60 | 0 | 1.000 (1.000-1.000) | 0.000 (0.000-0.000) | 0.000 (0.000-0.000) | 0.000 (0.000-0.000) | 0 | 0 | 0 | 60 |
| R54: Maintain: vedolizumab | recommended | 0 |  | NA | NA | NA | NA | 0 | 0 | 0 | 0 |
| R54: Maintain: vedolizumab | ordered | 0 |  | NA | NA | NA | NA | 0 | 0 | 0 | 0 |
| R54: Maintain: vedolizumab | executed | 0 |  | NA | NA | NA | NA | 0 | 0 | 0 | 0 |
| Overall | applies | 540 | 0.594 | 0.978 (0.969-0.985) | 0.982 (0.973-0.988) | 0.976 (0.972-0.982) | 0.988 (0.972-0.997) | 319 | 8 | 2 | 211 |
| Overall | recommended | 321 | 0.654 | 0.930 (0.925-0.935) | 0.947 (0.944-0.951) | 0.928 (0.927-0.932) | 0.968 (0.962-0.971) | 203 | 16 | 7 | 95 |
| Overall | ordered | 321 | 0.885 | 0.940 (0.935-0.947) | 0.966 (0.963-0.970) | 0.964 (0.961-0.968) | 0.968 (0.958-0.975) | 276 | 11 | 8 | 26 |
| Overall | executed | 321 | 0.863 | 0.923 (0.919-0.931) | 0.955 (0.953-0.960) | 0.960 (0.957-0.964) | 0.951 (0.946-0.957) | 264 | 11 | 13 | 33 |
| Overall | all (pooled) | 1503 | 0.727 | 0.948 (0.942-0.953) | 0.964 (0.960-0.968) | 0.959 (0.958-0.963) | 0.969 (0.962-0.975) | 1063 | 45 | 29 | 366 |

#### Supplementary Table 5: Proportion in eight states of adherence monitoring per guideline

For each of the guidelines the hospital days where a guideline item applied were grouped into eight categories denoted by R±O±E± shorthand indicating whether recommendation, ordered, and executed occurred. The percentage of hospital days in each category is denoted.

| Guideline Item | N | total applies | state_label | state_interpretation | % |
| --- | --- | --- | --- | --- | --- |
| R46: C. diff testing | 8 | 686 | R−O−E− | Broke at recognition | 1.2 |
| R46: C. diff testing | 21 | 686 | R−O−E+ | Executed w/o rec or order | 3.1 |
| R46: C. diff testing | 3 | 686 | R+O−E− | Broke at order | 0.4 |
| R46: C. diff testing | 7 | 686 | R+O−E+ | Executed without order | 1 |
| R46: C. diff testing | 23 | 686 | R+O+E− | Broke at execution | 3.4 |
| R46: C. diff testing | 624 | 686 | R+O+E+ | Full adherence | 91 |
| R47: DVT prophylaxis | 52 | 686 | R−O−E− | Broke at recognition | 7.6 |
| R47: DVT prophylaxis | 1 | 686 | R−O+E− | Ordered w/o rec, not executed | 0.1 |
| R47: DVT prophylaxis | 54 | 686 | R+O−E− | Broke at order | 7.9 |
| R47: DVT prophylaxis | 2 | 686 | R+O−E+ | Executed without order | 0.3 |
| R47: DVT prophylaxis | 111 | 686 | R+O+E− | Broke at execution | 16.2 |
| R47: DVT prophylaxis | 466 | 686 | R+O+E+ | Full adherence | 67.9 |
| R48: Avoid antibiotics | 26 | 686 | R−O−E− | Broke at recognition | 3.8 |
| R48: Avoid antibiotics | 9 | 686 | R−O−E+ | Executed w/o rec or order | 1.3 |
| R48: Avoid antibiotics | 188 | 686 | R−O+E+ | Silent adherence | 27.4 |
| R48: Avoid antibiotics | 10 | 686 | R+O−E− | Broke at order | 1.5 |
| R48: Avoid antibiotics | 7 | 686 | R+O−E+ | Executed without order | 1 |
| R48: Avoid antibiotics | 446 | 686 | R+O+E+ | Full adherence | 65 |
| R49: Avoid TPN | 3 | 686 | R−O−E− | Broke at recognition | 0.4 |
| R49: Avoid TPN | 682 | 686 | R−O+E+ | Silent adherence | 99.4 |
| R49: Avoid TPN | 1 | 686 | R+O+E+ | Full adherence | 0.1 |
| R50: IV steroids | 58 | 686 | R−O−E− | Broke at recognition | 8.5 |
| R50: IV steroids | 1 | 686 | R−O+E− | Ordered w/o rec, not executed | 0.1 |
| R50: IV steroids | 5 | 686 | R−O+E+ | Silent adherence | 0.7 |
| R50: IV steroids | 27 | 686 | R+O−E− | Broke at order | 3.9 |
| R50: IV steroids | 1 | 686 | R+O−E+ | Executed without order | 0.1 |
| R50: IV steroids | 17 | 686 | R+O+E− | Broke at execution | 2.5 |
| R50: IV steroids | 577 | 686 | R+O+E+ | Full adherence | 84.1 |
| R51: Rescue therapy day 3 | 35 | 273 | R−O−E− | Broke at recognition | 12.8 |
| R51: Rescue therapy day 3 | 1 | 273 | R−O−E+ | Executed w/o rec or order | 0.4 |
| R51: Rescue therapy day 3 | 13 | 273 | R+O−E− | Broke at order | 4.8 |
| R51: Rescue therapy day 3 | 2 | 273 | R+O−E+ | Executed without order | 0.7 |
| R51: Rescue therapy day 3 | 222 | 273 | R+O+E+ | Full adherence | 81.3 |
| R52: Maintain: IFX | 1 | 2 | R−O−E− | Broke at recognition | 50 |
| R52: Maintain: IFX | 1 | 2 | R+O−E− | Broke at order | 50 |

#### Supplementary Table 6: Cascade transition probabilities

For each guideline item the transition probability through the cascade is depicted. Confidence intervals are derived from Wilson. Guideline items R52-R54 are not shown as applicability was 0 for all.

| Guideline Item | Metric | k | n | Rate | CI_lower | CI_upper |
| --- | --- | --- | --- | --- | --- | --- |
| R46: C. diff testing | Recognition rate (P(Rec\|Applies)) | 657 | 686 | 0.958 | 0.94 | 0.97 |
| R46: C. diff testing | Order-follow-through (P(Ord\|Rec)) | 647 | 657 | 0.985 | 0.97 | 0.992 |
| R46: C. diff testing | Execution-given-order (P(Exe\|Ord)) | 624 | 647 | 0.964 | 0.95 | 0.976 |
| R46: C. diff testing | Silent execution (P(Exe\|Applies & ¬Rec)) | 21 | 29 | 0.724 | 0.54 | 0.853 |
| R46: C. diff testing | Overall execution (P(Exe\|Applies)) | 652 | 686 | 0.95 | 0.93 | 0.964 |
| R47: DVT prophylaxis | Recognition rate (P(Rec\|Applies)) | 633 | 686 | 0.923 | 0.9 | 0.94 |
| R47: DVT prophylaxis | Order-follow-through (P(Ord\|Rec)) | 577 | 633 | 0.912 | 0.89 | 0.931 |
| R47: DVT prophylaxis | Execution-given-order (P(Exe\|Ord)) | 466 | 578 | 0.806 | 0.77 | 0.836 |
| R47: DVT prophylaxis | Silent execution (P(Exe\|Applies & ¬Rec)) | 0 | 53 | 0 | 0 | 0.068 |
| R47: DVT prophylaxis | Overall execution (P(Exe\|Applies)) | 468 | 686 | 0.682 | 0.65 | 0.716 |
| R48: Avoid antibiotics | Recognition rate (P(Rec\|Applies)) | 463 | 686 | 0.675 | 0.64 | 0.709 |
| R48: Avoid antibiotics | Order-follow-through (P(Ord\|Rec)) | 446 | 463 | 0.963 | 0.94 | 0.977 |
| R48: Avoid antibiotics | Execution-given-order (P(Exe\|Ord)) | 634 | 634 | 1 | 0.99 | 1 |
| R48: Avoid antibiotics | Silent execution (P(Exe\|Applies & ¬Rec)) | 197 | 223 | 0.883 | 0.83 | 0.919 |
| R48: Avoid antibiotics | Overall execution (P(Exe\|Applies)) | 650 | 686 | 0.948 | 0.93 | 0.962 |
| R49: Avoid TPN | Recognition rate (P(Rec\|Applies)) | 1 | 686 | 0.001 | 0 | 0.008 |
| R49: Avoid TPN | Order-follow-through (P(Ord\|Rec)) | 1 | 1 | 1 | 0.21 | 1 |
| R49: Avoid TPN | Execution-given-order (P(Exe\|Ord)) | 683 | 683 | 1 | 0.99 | 1 |
| R49: Avoid TPN | Silent execution (P(Exe\|Applies & ¬Rec)) | 682 | 685 | 0.996 | 0.99 | 0.999 |
| R49: Avoid TPN | Overall execution (P(Exe\|Applies)) | 683 | 686 | 0.996 | 0.99 | 0.999 |
| R50: IV steroids | Recognition rate (P(Rec\|Applies)) | 622 | 686 | 0.907 | 0.88 | 0.926 |
| R50: IV steroids | Order-follow-through (P(Ord\|Rec)) | 594 | 622 | 0.955 | 0.94 | 0.969 |
| R50: IV steroids | Execution-given-order (P(Exe\|Ord)) | 582 | 600 | 0.97 | 0.95 | 0.981 |
| R50: IV steroids | Silent execution (P(Exe\|Applies & ¬Rec)) | 5 | 64 | 0.078 | 0.03 | 0.17 |
| R50: IV steroids | Overall execution (P(Exe\|Applies)) | 583 | 686 | 0.85 | 0.82 | 0.875 |
| R51: Rescue therapy day 3 | Recognition rate (P(Rec\|Applies)) | 237 | 273 | 0.868 | 0.82 | 0.903 |
| R51: Rescue therapy day 3 | Order-follow-through (P(Ord\|Rec)) | 222 | 237 | 0.937 | 0.9 | 0.961 |
| R51: Rescue therapy day 3 | Execution-given-order (P(Exe\|Ord)) | 222 | 222 | 1 | 0.98 | 1 |
| R51: Rescue therapy day 3 | Silent execution (P(Exe\|Applies & ¬Rec)) | 1 | 36 | 0.028 | 0 | 0.142 |
| R51: Rescue therapy day 3 | Overall execution (P(Exe\|Applies)) | 225 | 273 | 0.824 | 0.77 | 0.865 |
| R52: Maintain: IFX | Recognition rate (P(Rec\|Applies)) | 1 | 2 | 0.5 | 0.09 | 0.905 |
| R52: Maintain: IFX | Order-follow-through (P(Ord\|Rec)) | 0 | 1 | 0 | 0 | 0.793 |
| R52: Maintain: IFX | Execution-given-order (P(Exe\|Ord)) | 0 | 0 | 0 | 0 | 0 |
| R52: Maintain: IFX | Silent execution (P(Exe\|Applies & ¬Rec)) | 0 | 1 | 0 | 0 | 0.793 |
| R52: Maintain: IFX | Overall execution (P(Exe\|Applies)) | 0 | 2 | 0 | 0 | 0.658 |

#### Supplementary Table 7: Temporality of cascade steps

| Guideline Item | Days since admission | Gap label | n | n_total | rate |
| --- | --- | --- | --- | --- | --- |
| R46: C. diff testing | 1 | No recommendation (any R−) | 6 | 67 | 0.08955224 |
| R46: C. diff testing | 1 | No order (any O−) | 9 | 67 | 0.13432836 |
| R46: C. diff testing | 1 | No execution (any E−) | 20 | 67 | 0.29850746 |
| R46: C. diff testing | 2 | No recommendation (any R−) | 4 | 121 | 0.03305785 |
| R46: C. diff testing | 2 | No order (any O−) | 7 | 121 | 0.05785124 |
| R46: C. diff testing | 2 | No execution (any E−) | 4 | 121 | 0.03305785 |
| R46: C. diff testing | 3 | No recommendation (any R−) | 4 | 118 | 0.03389831 |
| R46: C. diff testing | 3 | No order (any O−) | 6 | 118 | 0.05084746 |
| R46: C. diff testing | 3 | No execution (any E−) | 3 | 118 | 0.02542373 |
| R46: C. diff testing | 4 | No recommendation (any R−) | 4 | 110 | 0.03636364 |
| R46: C. diff testing | 4 | No order (any O−) | 5 | 110 | 0.04545455 |
| R46: C. diff testing | 4 | No execution (any E−) | 2 | 110 | 0.01818182 |
| R46: C. diff testing | 5 | No recommendation (any R−) | 4 | 104 | 0.03846154 |
| R46: C. diff testing | 5 | No order (any O−) | 5 | 104 | 0.04807692 |
| R46: C. diff testing | 5 | No execution (any E−) | 2 | 104 | 0.01923077 |
| R46: C. diff testing | 6 | No recommendation (any R−) | 4 | 89 | 0.04494382 |
| R46: C. diff testing | 6 | No order (any O−) | 4 | 89 | 0.04494382 |
| R46: C. diff testing | 6 | No execution (any E−) | 1 | 89 | 0.01123596 |
| R46: C. diff testing | 7 | No recommendation (any R−) | 3 | 77 | 0.03896104 |
| R46: C. diff testing | 7 | No order (any O−) | 3 | 77 | 0.03896104 |
| R46: C. diff testing | 7 | No execution (any E−) | 2 | 77 | 0.02597403 |
| R47: DVT prophylaxis | 1 | No recommendation (any R−) | 16 | 67 | 0.23880597 |
| R47: DVT prophylaxis | 1 | No order (any O−) | 25 | 67 | 0.37313433 |
| R47: DVT prophylaxis | 1 | No execution (any E−) | 44 | 67 | 0.65671642 |
| R47: DVT prophylaxis | 2 | No recommendation (any R−) | 12 | 121 | 0.09917355 |
| R47: DVT prophylaxis | 2 | No order (any O−) | 24 | 121 | 0.19834711 |
| R47: DVT prophylaxis | 2 | No execution (any E−) | 45 | 121 | 0.37190083 |
| R47: DVT prophylaxis | 3 | No recommendation (any R−) | 12 | 118 | 0.10169492 |
| R47: DVT prophylaxis | 3 | No order (any O−) | 18 | 118 | 0.15254237 |
| R47: DVT prophylaxis | 3 | No execution (any E−) | 37 | 118 | 0.31355932 |
| R47: DVT prophylaxis | 4 | No recommendation (any R−) | 7 | 110 | 0.06363636 |
| R47: DVT prophylaxis | 4 | No order (any O−) | 13 | 110 | 0.11818182 |
| R47: DVT prophylaxis | 4 | No execution (any E−) | 31 | 110 | 0.28181818 |
| R47: DVT prophylaxis | 5 | No recommendation (any R−) | 2 | 104 | 0.01923077 |
| R47: DVT prophylaxis | 5 | No order (any O−) | 12 | 104 | 0.11538462 |
| R47: DVT prophylaxis | 5 | No execution (any E−) | 26 | 104 | 0.25 |
| R47: DVT prophylaxis | 6 | No recommendation (any R−) | 2 | 89 | 0.02247191 |
| R47: DVT prophylaxis | 6 | No order (any O−) | 10 | 89 | 0.11235955 |
| R47: DVT prophylaxis | 6 | No execution (any E−) | 19 | 89 | 0.21348315 |
| R47: DVT prophylaxis | 7 | No recommendation (any R−) | 2 | 77 | 0.02597403 |
| R47: DVT prophylaxis | 7 | No order (any O−) | 6 | 77 | 0.07792208 |
| R47: DVT prophylaxis | 7 | No execution (any E−) | 16 | 77 | 0.20779221 |
| R48: Avoid antibiotics | 1 | No recommendation (any R−) | 53 | 67 | 0.79104478 |
| R48: Avoid antibiotics | 1 | No order (any O−) | 6 | 67 | 0.08955224 |
| R48: Avoid antibiotics | 1 | No execution (any E−) | 5 | 67 | 0.07462687 |
| R48: Avoid antibiotics | 2 | No recommendation (any R−) | 42 | 121 | 0.34710744 |
| R48: Avoid antibiotics | 2 | No order (any O−) | 10 | 121 | 0.08264463 |
| R48: Avoid antibiotics | 2 | No execution (any E−) | 7 | 121 | 0.05785124 |
| R48: Avoid antibiotics | 3 | No recommendation (any R−) | 37 | 118 | 0.31355932 |
| R48: Avoid antibiotics | 3 | No order (any O−) | 13 | 118 | 0.11016949 |
| R48: Avoid antibiotics | 3 | No execution (any E−) | 8 | 118 | 0.06779661 |
| R48: Avoid antibiotics | 4 | No recommendation (any R−) | 29 | 110 | 0.26363636 |
| R48: Avoid antibiotics | 4 | No order (any O−) | 7 | 110 | 0.06363636 |
| R48: Avoid antibiotics | 4 | No execution (any E−) | 4 | 110 | 0.03636364 |
| R48: Avoid antibiotics | 5 | No recommendation (any R−) | 24 | 104 | 0.23076923 |
| R48: Avoid antibiotics | 5 | No order (any O−) | 7 | 104 | 0.06730769 |
| R48: Avoid antibiotics | 5 | No execution (any E−) | 4 | 104 | 0.03846154 |
| R48: Avoid antibiotics | 6 | No recommendation (any R−) | 20 | 89 | 0.2247191 |
| R48: Avoid antibiotics | 6 | No order (any O−) | 5 | 89 | 0.05617978 |
| R48: Avoid antibiotics | 6 | No execution (any E−) | 4 | 89 | 0.04494382 |
| R48: Avoid antibiotics | 7 | No recommendation (any R−) | 18 | 77 | 0.23376623 |
| R48: Avoid antibiotics | 7 | No order (any O−) | 4 | 77 | 0.05194805 |
| R48: Avoid antibiotics | 7 | No execution (any E−) | 4 | 77 | 0.05194805 |
| R49: Avoid TPN | 1 | No recommendation (any R−) | 67 | 67 | 1 |
| R49: Avoid TPN | 1 | No order (any O−) | 0 | 67 | 0 |
| R49: Avoid TPN | 1 | No execution (any E−) | 0 | 67 | 0 |
| R49: Avoid TPN | 2 | No recommendation (any R−) | 120 | 121 | 0.99173554 |
| R49: Avoid TPN | 2 | No order (any O−) | 0 | 121 | 0 |
| R49: Avoid TPN | 2 | No execution (any E−) | 0 | 121 | 0 |
| R49: Avoid TPN | 3 | No recommendation (any R−) | 118 | 118 | 1 |
| R49: Avoid TPN | 3 | No order (any O−) | 0 | 118 | 0 |
| R49: Avoid TPN | 3 | No execution (any E−) | 0 | 118 | 0 |
| R49: Avoid TPN | 4 | No recommendation (any R−) | 110 | 110 | 1 |
| R49: Avoid TPN | 4 | No order (any O−) | 1 | 110 | 0.00909091 |
| R49: Avoid TPN | 4 | No execution (any E−) | 1 | 110 | 0.00909091 |
| R49: Avoid TPN | 5 | No recommendation (any R−) | 104 | 104 | 1 |
| R49: Avoid TPN | 5 | No order (any O−) | 1 | 104 | 0.00961538 |
| R49: Avoid TPN | 5 | No execution (any E−) | 1 | 104 | 0.00961538 |
| R49: Avoid TPN | 6 | No recommendation (any R−) | 89 | 89 | 1 |
| R49: Avoid TPN | 6 | No order (any O−) | 0 | 89 | 0 |
| R49: Avoid TPN | 6 | No execution (any E−) | 0 | 89 | 0 |
| R49: Avoid TPN | 7 | No recommendation (any R−) | 77 | 77 | 1 |
| R49: Avoid TPN | 7 | No order (any O−) | 1 | 77 | 0.01298701 |
| R49: Avoid TPN | 7 | No execution (any E−) | 1 | 77 | 0.01298701 |
| R50: IV steroids | 1 | No recommendation (any R−) | 20 | 67 | 0.29850746 |
| R50: IV steroids | 1 | No order (any O−) | 36 | 67 | 0.53731343 |
| R50: IV steroids | 1 | No execution (any E−) | 47 | 67 | 0.70149254 |
| R50: IV steroids | 2 | No recommendation (any R−) | 16 | 121 | 0.1322314 |
| R50: IV steroids | 2 | No order (any O−) | 20 | 121 | 0.16528926 |
| R50: IV steroids | 2 | No execution (any E−) | 24 | 121 | 0.19834711 |
| R50: IV steroids | 3 | No recommendation (any R−) | 11 | 118 | 0.09322034 |
| R50: IV steroids | 3 | No order (any O−) | 13 | 118 | 0.11016949 |
| R50: IV steroids | 3 | No execution (any E−) | 14 | 118 | 0.11864407 |
| R50: IV steroids | 4 | No recommendation (any R−) | 8 | 110 | 0.07272727 |
| R50: IV steroids | 4 | No order (any O−) | 8 | 110 | 0.07272727 |
| R50: IV steroids | 4 | No execution (any E−) | 8 | 110 | 0.07272727 |
| R50: IV steroids | 5 | No recommendation (any R−) | 5 | 104 | 0.04807692 |
| R50: IV steroids | 5 | No order (any O−) | 4 | 104 | 0.03846154 |
| R50: IV steroids | 5 | No execution (any E−) | 5 | 104 | 0.04807692 |
| R50: IV steroids | 6 | No recommendation (any R−) | 2 | 89 | 0.02247191 |
| R50: IV steroids | 6 | No order (any O−) | 3 | 89 | 0.03370787 |
| R50: IV steroids | 6 | No execution (any E−) | 2 | 89 | 0.02247191 |
| R50: IV steroids | 7 | No recommendation (any R−) | 2 | 77 | 0.02597403 |
| R50: IV steroids | 7 | No order (any O−) | 2 | 77 | 0.02597403 |
| R50: IV steroids | 7 | No execution (any E−) | 3 | 77 | 0.03896104 |
| R51: Rescue therapy day 3 | 1 | No recommendation (any R−) | 1 | 1 | 1 |
| R51: Rescue therapy day 3 | 1 | No order (any O−) | 1 | 1 | 1 |
| R51: Rescue therapy day 3 | 1 | No execution (any E−) | 1 | 1 | 1 |
| R51: Rescue therapy day 3 | 2 | No recommendation (any R−) | 2 | 18 | 0.11111111 |
| R51: Rescue therapy day 3 | 2 | No order (any O−) | 3 | 18 | 0.16666667 |
| R51: Rescue therapy day 3 | 2 | No execution (any E−) | 2 | 18 | 0.11111111 |
| R51: Rescue therapy day 3 | 3 | No recommendation (any R−) | 0 | 28 | 0 |
| R51: Rescue therapy day 3 | 3 | No order (any O−) | 0 | 28 | 0 |
| R51: Rescue therapy day 3 | 3 | No execution (any E−) | 0 | 28 | 0 |
| R51: Rescue therapy day 3 | 4 | No recommendation (any R−) | 6 | 46 | 0.13043478 |
| R51: Rescue therapy day 3 | 4 | No order (any O−) | 9 | 46 | 0.19565217 |
| R51: Rescue therapy day 3 | 4 | No execution (any E−) | 8 | 46 | 0.17391304 |
| R51: Rescue therapy day 3 | 5 | No recommendation (any R−) | 10 | 62 | 0.16129032 |
| R51: Rescue therapy day 3 | 5 | No order (any O−) | 15 | 62 | 0.24193548 |
| R51: Rescue therapy day 3 | 5 | No execution (any E−) | 15 | 62 | 0.24193548 |
| R51: Rescue therapy day 3 | 6 | No recommendation (any R−) | 9 | 64 | 0.140625 |
| R51: Rescue therapy day 3 | 6 | No order (any O−) | 15 | 64 | 0.234375 |
| R51: Rescue therapy day 3 | 6 | No execution (any E−) | 14 | 64 | 0.21875 |
| R51: Rescue therapy day 3 | 7 | No recommendation (any R−) | 8 | 54 | 0.14814815 |
| R51: Rescue therapy day 3 | 7 | No order (any O−) | 8 | 54 | 0.14814815 |
| R51: Rescue therapy day 3 | 7 | No execution (any E−) | 8 | 54 | 0.14814815 |

#### Supplementary Table 8: Trend analysis of execution gaps over time

Spearman correlation of year and rate of execution gaps per category and overall.

| **Guideline Item** | **Spearman Correlation** | **p-value** |
| --- | --- | --- |
| overall | -0.428571 | 0.396501 |
| R46: C. diff testing | -0.314286 | 0.544093 |
| R47: DVT prophylaxis | -0.428571 | 0.396501 |
| R48: Avoid antibiotics | 0.657143 | 0.156175 |
| R49: Avoid TPN | 0.371868 | 0.467910 |
| R50: IV steroids | -0.771429 | 0.072397 |
| R51: Rescue therapy day 3 | 0.542857 | 0.265703 |
